# A germline heterozygous dominant negative *IKZF2* variant causing syndromic primary immune regulatory disorder and ICHAD

**DOI:** 10.1101/2023.09.09.23295301

**Authors:** Henry Y. Lu, Maryam Vaseghi-Shanjani, Avery J. Lam, Mehul Sharma, Arezoo Mohajeri, Jana Gillies, Gui Xiang Yang, Susan Lin, Maggie P. Fu, Areesha Salman, Ronak Rahmanian, Linlea Armstrong, Jessica Halparin, Connie L. Yang, Mark Chilvers, Erika Henkelman, Wingfield Rehmus, Douglas Morrison, Audi Setiadi, Sara Mostafavi, Michael S. Kobor, Frederick K. Kozak, Catherine M. Biggs, Clara van Karnebeek, Kyla J. Hildebrand, Anna Lehman the Care4Rare Canada Consortium, Megan K. Levings, Stuart E. Turvey

## Abstract

Monogenic defects that impair the control of inflammation and tolerance lead to profound immune dysregulation, including autoimmunity and atopy. Studying these disorders reveals important molecular and cellular factors that regulate human immune homeostasis and identifies potential precision medicine targets. Here, we provide a detailed immunological assessment of a pediatric patient with a recently discovered syndrome causing Immunodysregulation, Craniofacial anomalies, Hearing impairment, Athelia, and Developmental delay (or ICHAD syndrome). The immunodysregulation resulted in autoimmune hemolytic anemia (AIHA) and atopic dermatitis. The patient carried a *de novo* germline heterozygous c.406+540_574+13477dup;p.Gly136_Ser191dup variant in *IKAROS family zinc finger 2* (*IKZF2*), which encodes Helios. This variant led to reduced Helios protein expression and dominant interference of wild-type Helios-mediated repression of the *IL2* promoter. Multi-parameter flow cytometric analyses of patient peripheral blood mononuclear cells revealed strongly impaired natural killer cell differentiation and function, and increased CD8^+^ T cell activation and cytokine secretion. Strikingly, patient CD4^+^ T cells were hyperactive, produced elevated levels of nearly all T helper (T_H_) cytokines, and readily proliferated in response to stimulation. Patient regulatory T cells (Tregs) developed normally but aberrantly produced high levels of many T_H_ cytokines. Single-cell RNA sequencing revealed largely normal Tregs (albeit mostly memory), but naïve CD4^+^ T cells that were more enriched in genes related to activation, proliferation, metabolism, and T_H_ differentiation. This work describes the immunological phenotype of one of the first reported cases of germline dominant negative Helios deficiency, expands our understanding of the pathogenesis of AIHA on a single cell level, and provides valuable insights into Helios function in a variety of lymphocyte subsets.

## INTRODUCTION

Immunity is tightly regulated to optimize protective responses against pathogens and malignancy while preventing inappropriate responses to otherwise benign antigens. Monogenic defects that impair this regulation of inflammation or tolerance result in a subgroup of the inborn errors of immunity (IEI) called primary immune regulatory disorders (PIRD)^1^. In contrast to classic IEIs, which typically manifest as unusually severe or recurrent infections, PIRDs predominantly present with immune-mediated pathology, including autoimmunity, lymphoproliferation, malignancy, autoinflammation, and atopy, with susceptibility to infections being a less pronounced aspect of these disorders^2^.

Immune dysregulation can lead to a variety of hematologic manifestations and cytopenias^3^. A classic example is autoimmune hemolytic anemia (AIHA) caused by immune-mediated destruction of erythrocytes^4^. AIHA can be idiopathic (primary) or secondary and is classified as warm or cold (cold agglutinin disease [CAD], paroxysmal cold hemoglobinuria) depending on autoantibody behaviour at different thermal ranges, which is important for determining appropriate treatment regimens^5^. Most cases of AIHA are considered idiopathic (∼60%)^6^, while secondary causes can include IEIs/PIRDs, malignancy, bacterial/viral infections, and drugs^4^. As such, the etiology and pathogenesis of AIHA remains incompletely understood.

In the past 2 years, human germline variants in a family of transcription factors called the IKAROS zinc finger (IKZF) family have been linked to immunodeficiency and cytopenias^7–10^. Notably, loss-of-function (LOF) variants in *IKZF2* (Helios) were recently associated with combined immunodeficiency (CID) and/or immune dysregulation, including immune thrombocytopenia (ITP), Evan’s syndrome, and systemic lupus erythematosus^7–9^. *IKZF2* is highly expressed in hematopoietic stem/progenitor cells, T cells, including activated CD4^+^/CD8^+^ T cells, mucosa-associated invariant T (MAIT) cells, regulatory T cells (Tregs), and natural killer (NK) cells^11–16^. As Tregs are among the highest *IKZF2* expressors, most studies have focused on the role of Helios in Treg development and function^17–27^. In mice, Helios is critical for Treg identity, survival, and stability, but is seemingly dispensable in mice and humans for suppressive function^20,27^.

Nevertheless, both *IKZF2^-/-^* and Treg-specific *IKZF2^-/-^*mice develop progressive autoimmune disease associated with activated CD4^+^/CD8^+^ T cells, increased T follicular helper (T_FH_) and germinal center (GC) B cell numbers, autoantibody production, and increased proinflammatory cytokine production^19,20,28^. The role of *IKZF2* in other components of the human immune system (e.g. NK cells) is beginning to be better defined through the identification of humans with germline *IKZF2* variants^7–10^.

Here, we report a detailed immunological and mechanistic workup of the first reported case of germline heterozygous dominant negative (DN) *IKZF2* disorder in a young girl we recently described^10^. This patient was found to have a novel genetic syndrome comprising Immunodysregulation, Craniofacial anomalies, Hearing impairment, Athelia, and Developmental delay (or ICHAD syndrome). We report a comprehensive mechanistic immune evaluation of this patient which revealed significant developmental and functional defects in NK and naïve CD4^+^ T cells and marked immune activation as the likely cause of immune dysregulation. Informed by our improved appreciation of the human phenotype caused by germline IKZF2 variants, we suggest that germline heterozygous dominant negative *IKZF2* variants should now be considered in the differential diagnosis of patients with AIHA and PIRD.

## METHODS

### Study participants and consent

All study participants and/or their parents/guardians provided written informed consent. Research study protocols (H18-02853, H18-02912, H15-00092) were approved by The University of British Columbia Clinical Research Ethics Board.

### Cell isolation, culture, and immortalization

Peripheral blood mononuclear cells (PBMCs) were isolated from all study participants by standard Ficoll-Paque (GE Healthcare) density centrifugation as previously described^29^. Lymphoblastoid cell lines (LCLs) were derived by standard EBV transformation as previously described^29^ (**Supplement**).

### Plasmid generation and cloning

WT *IZKF2*, p.Gly136_Ser191dup *IKZF2*, pIL2-Luc2 plasmids were generated by introducing restriction sites into cDNA or genomic DNA by PCR and cloning into pCMV6-XL4-3xFLAG and p.GL4.14 firefly luciferase gene reporter (Promega) vectors, respectively (**Supplement**).

### Transient transfections

HEK293 cells were transfected with empty vector (EV), WT, p.Gly136_Ser191dup, or varying ratios of WT/p.Gly136_Ser191dup *IKZF2* expression plasmids by Lipofectamine 3000 (ThermoFisher) transfection (**Supplement**). Transfected cells were then processed for immunoblotting or immunofluorescence. For luciferase assays, cells were additionally transfected with pIL2-Luc2 and Renilla luciferase (R-Luc) plasmids then processed with a Dual-Glo Luciferase Assay Kit (#E2920, Promega) (**Supplement**).

### Immunoblotting

Helios protein detection was accomplished in LCLs and transfected HEK293 cells by standard immunoblotting as previously described^29^ (**Supplement**).

### Immunofluorescence

12mm coverslips were sterilized, HEK293 cells were seeded on top in 6-well plates, allowed to attach for 24h, then transfected as above. Transfected cells were fixed, permeabilized, and stained for Helios, DAPI, and F-actin before mounting and imaging on a Leica SP5 II Laser Scanning Confocal Microscope (Leica Microsystems GmbH) (**Supplement**).

### Immunophenotyping

Immunophenotyping and intracellular cytokine was performed as previously described^30^. Briefly, PBMCs were stimulated with PMA and ionomycin (P/I) in the presence of GolgiStop (#554724, BD Biosciences), stained with antibody panels 1-4 (**Supplemental Table 5**) using the eBioscience Foxp3 Transcription Factor Staining Buffer Set (#00-5523-00, Invitrogen, ThermoFisher), acquired on a FACSymphony flow cytometer, and analyzed using FlowJo (both BD Biosciences) (**Supplement**).

### Proliferation assays

PBMCs were seeded at varying concentrations, labelled with Cell Proliferation Dye (CPD) eF450 (#65-0842-85, Invitrogen, ThermoFisher) and stimulated with anti-CD3/CD28-coated Dynabeads (#11141D, Human T-Expander, Gibco, ThermoFisher) for four days. Cells were stained with antibody panel 6 (**Supplemental Table 5**) and acquired on a CytoFLEX (Beckman Coulter) and analyzed using FlowJo (**Supplement**).

### T cell suppression assays

Treg suppression of CD3^+^ T cell proliferation was assessed as previously described^27^. Briefly, patient and control PBMCs were enriched for CD4^+^ T cells, sorted using panel 7 (**Supplemental Table 5**) for Tregs and Tconvs, expanded with IL-2 and irradiated mouse L cells for seven days, then cocultured with various ratios of CPD-labelled anti-CD3/CD28 Dynabead-stimulated enriched CD3^+^ T responder (Tresp) cells for four days. Percent suppression of CD4^+^ and CD8^+^ T cell proliferation was calculated using division index (DI): (1 -[DI of sample/DI of positive control]) x 100% (**Supplement**).

### Quantification of cytokine production

Supernatants were collected from PBMCs stimulated for proliferation, cocultured Tregs and Tresp for T cell suppression assays, and stimulated Tregs and Tconvs alone. Cytokine concentrations were measured by LEGENDplex Human Th Cytokine Panel 12-plex (#741027, BioLegend) on a CytoFLEX (Beckman Coulter) and analyzed with Qognit software (BioLegend) according to manufacturer’s recommendations (**Supplement**).

### TSDR methylation

Genomic DNA was isolated from expanded Tregs and Tconvs, bisulfite converted with an EZ DNA Methylation-Direct Kit (Zymo Research), PCR amplified, and pyrosequenced using a PyroMark Q96 MD (Qiagen) as previously described^27^ (**Supplement**).

### Single-cell RNA sequencing

Single-cell RNA sequencing was performed on unstimulated and 4h P/I-stimulated sorted Tregs and Tconv from P1 and two age-matched/sex-matched controls using the BD Rhapsody Single Cell platform (BD Biosciences) according to manufacturer’s recommendations and analyzed as previously described^30^ (**Supplement**). Raw data are deposited on Gene Expression Omnibus.

## RESULTS

### Clinical case presentation

The index patient (P1) is a 5-10-year-old girl born to non-consanguineous parents (**Figure 1A**) who presented with syndromic features of developmental abnormalities and immune dysregulation. Family history is unremarkable. She has profound bilateral sensorineural hearing loss, microcephaly, mild developmental delay, cleft palate, hypotonia, athelia, and dysmorphic facies (detailed in^10^). Consistent with underlying immune dysregulation, she has suffered from chronic anemia beginning at two months of age, which at times required hospitalization and red blood cell transfusions (**Figure 1B-I**). She also had severe atopic dermatitis beginning at ten months of age affecting the face, arms, legs, and trunk. Her skin inflammation was initially refractory to optimal medical therapy (i.e. topical corticosteroids, emollients, bleach baths), but has become more manageable with age. The patient also experienced frequent upper respiratory tract infections, which reduced in frequency following initiation of monthly intravenous immunoglobulin (IVIG) replacement therapy. Laboratory and hematologic evaluation revealed chronically positive direct antiglobulin tests (DAT) initially consistent with a mixed (cold>warm) AIHA phenotype, which progressed to a predominantly warm AIHA phenotype with age without overt hemolysis. Other notable hematological features included a normocellular bone marrow with scattered hemophagocytic macrophages, no morphological evidence for dysplastic cells or a progressive marrow disorder, lymphopenia, and elevated IgE (**Table 1**). Red cells did not carry any hemoglobin variants, although non-specific poikilocytosis and rouleaux formation were noted.

**Figure 1.**
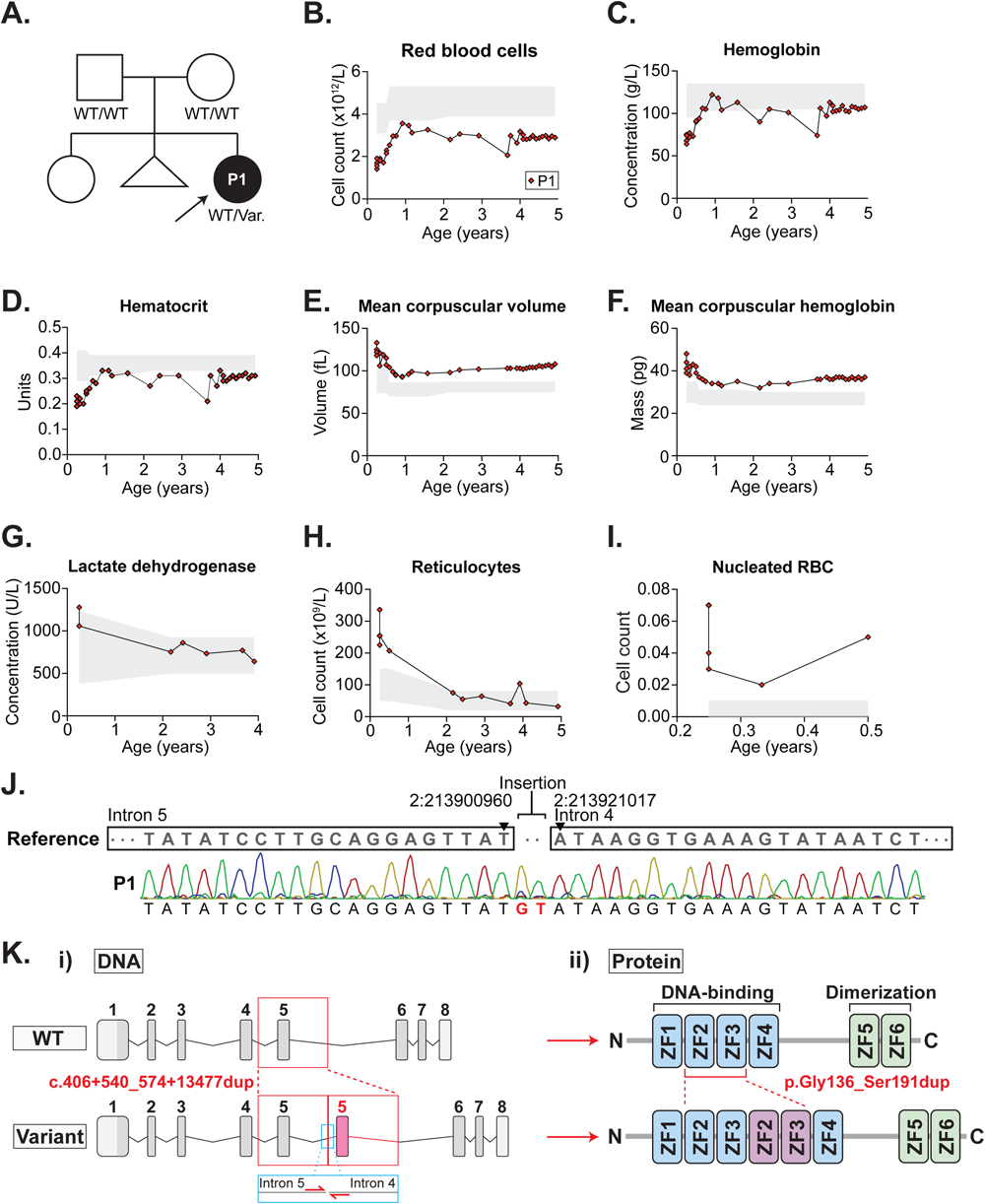
Clinical phenotype of the patient. A) Family pedigree of the patient. Filled symbols indicate affected individuals. Arrow represents index patient. Known genotype is marked. B-I) Hematological parameters measured in the patient over time. Shaded regions=age-specific reference ranges. J) Sanger sequencing of DNA extracted from whole blood of the patient. Primers were designed to flank the breakpoint. K) Schematic representation of the impact of i) the c.406+540_574+13477dup *IKZF2* variant on the genomic level. Red inset=exon 5 duplication. Blue inset=Sanger sequencing primer location; and ii) the p.Gly136_Ser191dup Helios variant on the protein level. Red inset=zinc finger 2 and 3 duplication. Var., variant; WT, wild-type; ZF, zinc finger; P1, patient 1.

**Table 1.**
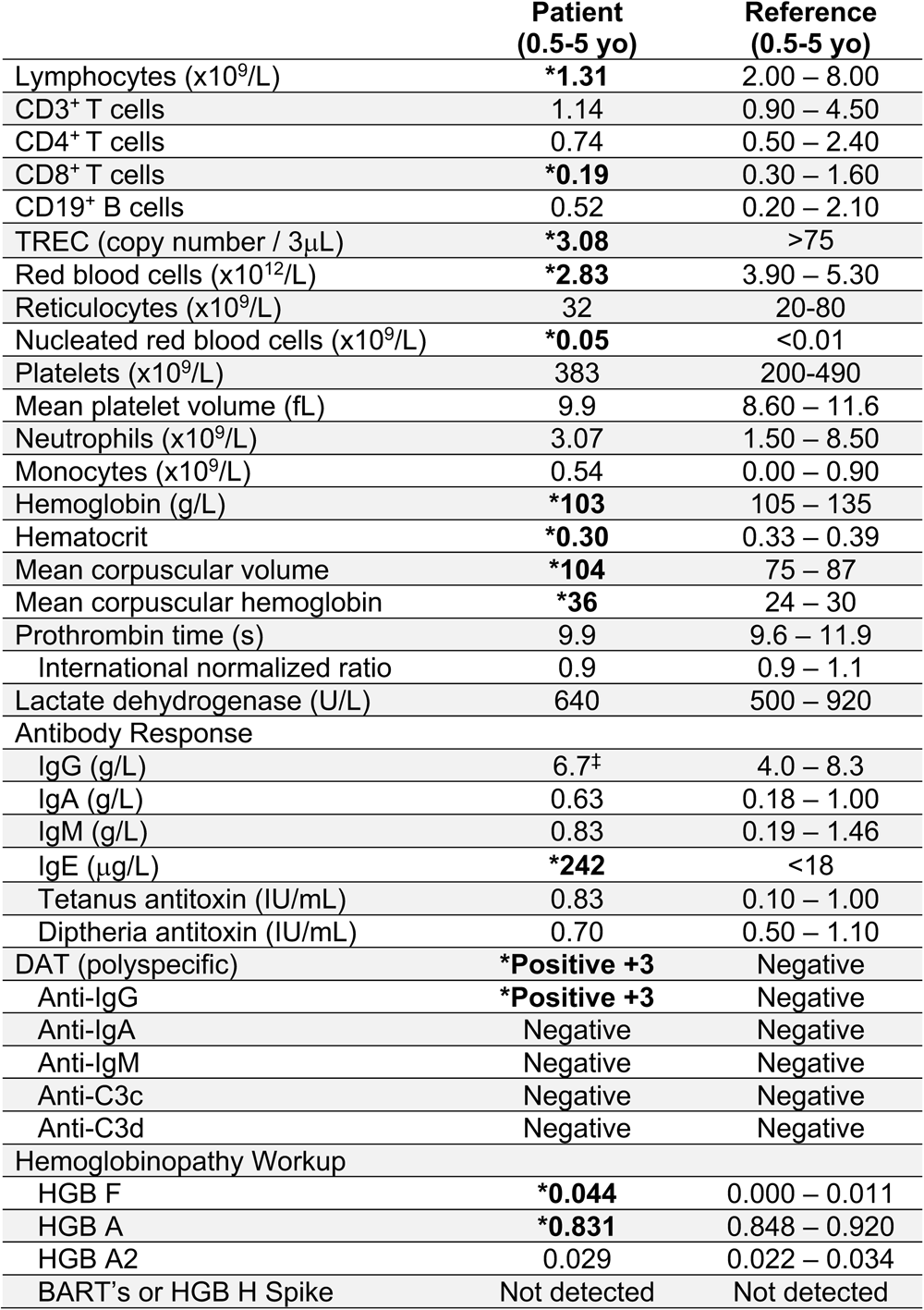
Summary of major hematological and immunological laboratory parameters. Tabulation of patient hematological and immunological laboratory values compared to age-specific reference ranges. ^‡^value measured while patient was on intravenous immunoglobulin replacement. TREC, T cell receptor excision circle; DAT, direct antiglobulin test; HGB, hemoglobin.

Given the unique constellation of features, P1 underwent trio WGS (detailed in^10^). This revealed a novel *de novo* germline heterozygous structural variant affecting *IKAROS zinc finger 2* (*IKZF2*) encoding Helios (NM_016260.3:c.406+540_574+13477dup; NP_057344.2:p.Gly136_Ser191dup) (**Figure 1J-K**). The variant results in intron 5 at chr2:213900960 being joined to intron 6 chr2:213921017 (GRCh37) with a 2 bp GT insertion in between. This leads to tandem duplication of exon 5 (∼20kb duplication spanning exon 5 and parts of flanking introns), corresponding to tandem duplication of zinc fingers 2 and 3 of Helios (**Figure 1K**).

### Characterization of the impact of the p.Gly136_Ser191dup Helios variant on protein expression and localization

The p.Gly136_Ser191dup Helios variant created a higher molecular weight Helios protein (as anticipated with a ∼13kDa 56 amino acid duplication) detected in both transfected HEK293 cells and primary patient-derived LCLs (**Figure 2A, C** and detailed in^10^). Total Helios protein expression was reduced overall in both transfected HEK293 (**Figure 2A-B**) and patient-derived LCLs (**Figure 2C-E**), likely because the variant protein was less stable. As a TF, Helios localizes to the nucleus to regulate transcription^12^. We transfected HEK293 cells with WT or p.Gly136_Ser191dup *IKZF2* alone, or increasing quantities of p.Gly136_Ser191dup together with WT *IKZF2* (**Figure 2F**) to assess protein expression by immunofluorescence. The variant protein was indistinguishable from WT protein in nuclear localization, nor did it affect the nuclear localization of WT protein.

**Figure 2.**
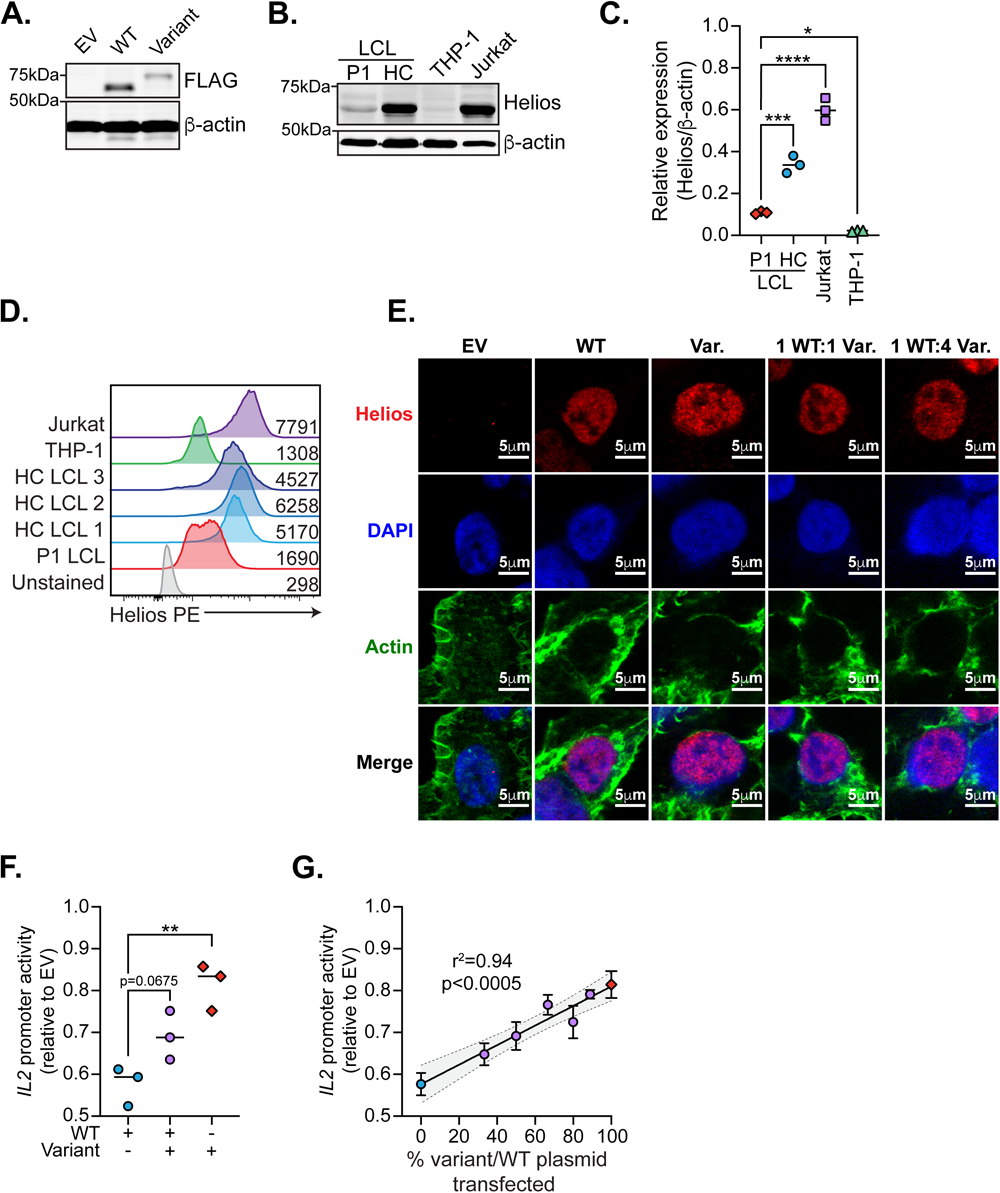
A novel *IKZF2* variant leads to reduced Helios protein expression and dominant interference of WT function. A-B) HEK293 cells were transfected with empty vector (EV), wild-type (WT) *IKZF2*, or p.Gly136_Ser191dup (variant) *IKZF2* alone or in different combinations. Expression was determined by immunoblotting with both FLAG and Helios. n=5. C-E) Expression of Helios was detected in patient-derived and control lymphoblastoid cell lines (LCLs) and compared to negative (THP-1 monocytic cells) and positive (Jurkat T leukemia cells) controls by C-D) immunoblot and E) flow cytometry. F) HEK293 cells were transfected with EV, WT, variant, or ratios of WT:variant and subjected to immunofluorescence for detection of Helios (red, Alexa Fluor 647), the nucleus (blue, DAPI), and F-actin (green, Phalloidin Alexa Fluor 488) using a Leica SP5 Confocal and LAS X Software with a 100x objective lens. Scale bars=5μm. n=3. G-H) HEK293 cells were cotransfected with EV, *IKZF2* expression plasmids at various ratios, pIL2-luc2, and Renilla luciferase. *IL2* promoter activity was defined as relative light units from different conditions normalized to EV. H) Pearson correlation between *IL2* promoter activity and % variant/WT *IKZF2* plasmid transfected. Shaded region=95% confidence interval. n=3. *p<0.05, **p<0.01, ***p<0.001, ****p<0.0001. Ordinary one-way ANOVA with Šidák’s multiple comparisons test.

### p.Gly136_Val192dup Helios acts through dominant interference

Helios has previously been shown to bind the *IL2* promoter to silence IL-2 production in Tregs^32^. We therefore cloned the human *IL2* promoter (-580 bp to +57 bp) into a firefly luciferase gene reporter vector and cotransfected this with Renilla luciferase, WT or p.Gly136_Ser191dup *IKZF2* alone, or p.Gly136_Ser191dup together with WT (**Figure 2G** and detailed in^10^). As expected, WT *IKZF2* strongly repressed *IL2* promoter activity. However, p.Gly136_Val192dup not only showed impaired repression on its own, but significantly interfered with WT *IKZF2*-mediated *IL2* repression (**Figure 2G**). This DN effect was dose-dependent, with increasing quantities of p.Gly136_Ser191dup transfected correlating with more impaired WT *IKZF2*-dependent *IL2* repression (r^2^=0.94, p<0.0005) (**Figure 2H** and detailed in^10^).

### Patient lymphocyte phenotyping reveals NK cell lymphocytosis and CD3^+^ T cell lymphopenia

Helios is highly expressed in CD3^+^ T cells and NK cells^11^, both of which are known to be dysregulated in AIHA^4^. We profiled the frequencies of CD3^+^ T, CD3^-^CD56^+^ NK, and CD3^+^CD56^+^ NKT cells as well as Helios expression from P1 (n=5 independent blood draws) and a cohort of adult (n=8) and age-matched controls (n=11). P1 had significantly reduced frequencies of CD3^+^ T (**Figure 3A-B**), markedly elevated NK (**Figure 3A,C**), and modestly elevated NKT (**Figure 3A,D**) cells, all of which were Helios-deficient (**Figure 3E-H**).

**Figure 3.**
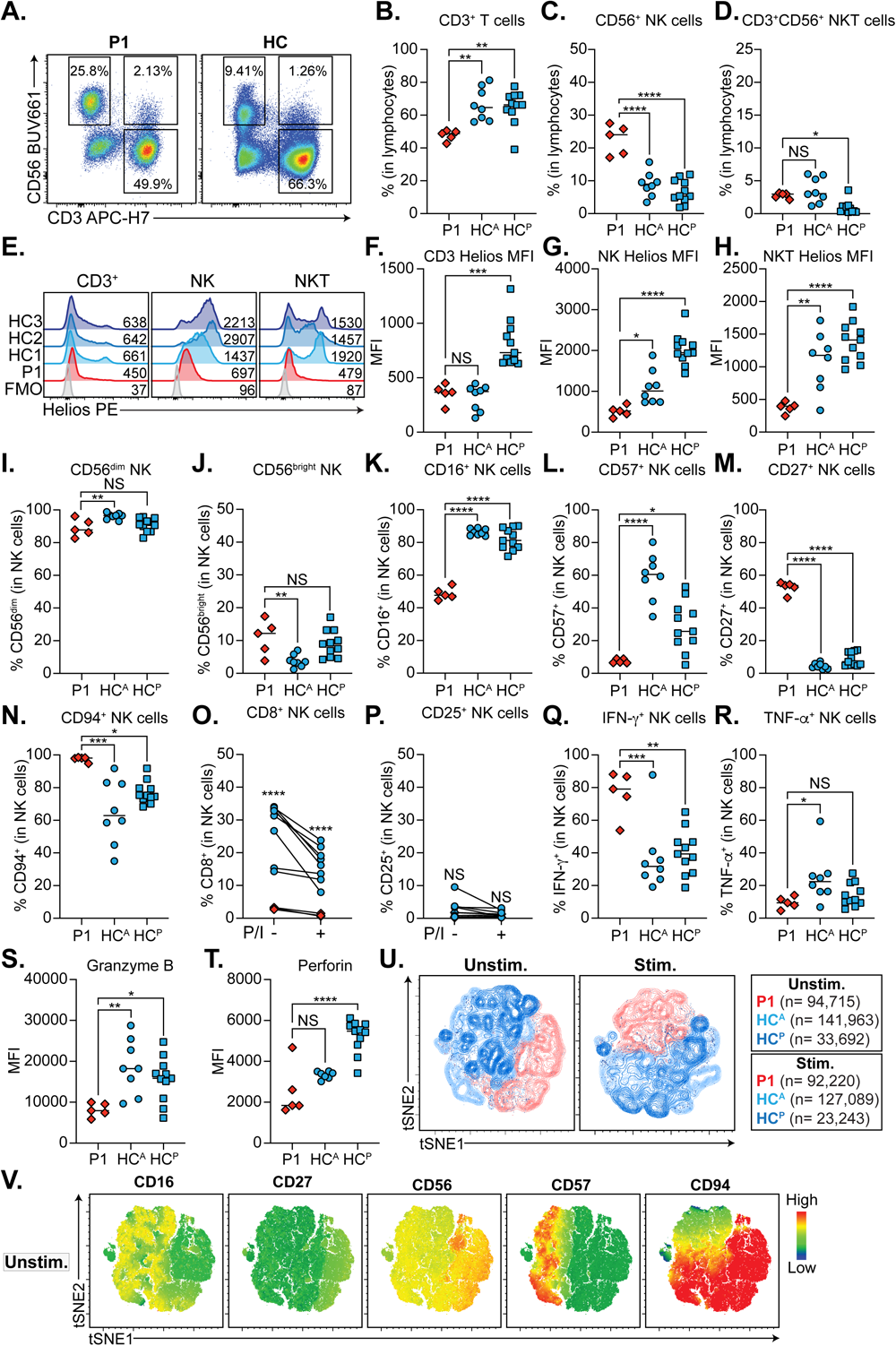
P1 has impaired NK cell development and function. A) Representative flow cytometry dot plots showing CD3^+^CD56^-^ T cells, CD3^-^CD56^+^ NK cells, and CD3^+^CD56^+^ NKT cells for P1 and a control. B-D) Frequency of B) T cells, C) NK cells, D) NKT cells from A) in P1 and adult (HC^A^) and pediatric (HC^P^) controls. E) Representative Helios histograms for T, NK, and NKT cells in P1 and controls compared to a fluorescence minus one (FMO) control. Mean fluorescence intensities (MFI) are indicated. F-H) Quantification of Helios MFI in F) T cells, G) NK cells, H) NKT cells from P1 and controls. I-N) Frequency of I) CD56^dim^, J) CD56^bright^ NK, K) CD16^+^, L) CD57^+^, M) CD27^+^, N) CD94^+^ NK cells in P1 compared to controls. O-P) Frequency of N) CD8^+^ and P) CD25^+^ NK cells in patient and controls before and after 4h PMA and ionomycin (P/I) stimulation. Q-R) Frequency of Q) IFN-γ^+^ and R) TNF-α^+^ NK cells in P1 and controls after P/I stimulation. S-T) MFI of S) granzyme B and T) perforin in unstimulated P1 and control NK cells. U-V) Live NK cells from P1 or controls clustered on CD16, CD27, CD57, CD94, Helios, perforin, granzyme B, IFN-γ, and TNF-α. Number of NK cells clustered for each group is indicated on the panels to the right. V) tSNE plots from U) coloured based on CD16, CD27, CD56, CD57, or CD94 expression. A-N, Q-T) P1 n=5, HC^A^ n=8, HC^P^ n=11. O-P) P1 n=3, HC n=8. *p<0.05, **p<0.01, ***p<0.001, ****p<0.0001. Ordinary one-way ANOVA with Šidák’s multiple comparisons test.

### P1 NK cells are phenotypically immature and functionally abnormal

Given the intriguing NK cell lymphocytosis, we set out to define the developmental and functional status of P1 NK cells by assessing the expression of CD56, maturation (CD27, CD57, CD94), adhesion (CD16), and activation (CD8, CD25) markers^33–35^ on P1 NK cells compared to controls. The relative abundance of CD56^dim^ and CD56^bright^ NK cell populations was normal (**Figure 3I-J**), although on histograms, the patient lacked a clear CD56^bright^ population (**Figure S1A**). Strikingly, all maturation markers assessed were dysregulated in P1, including significantly reduced CD16^+^ (**Figure 3K**, **Figure S1B**) and CD57^+^ (**Figure 3L**, **Figure S1C**), but elevated CD27^+^ (**Figure 3M**, **Figure S1D**) and CD94^+^ (**Figure 3N**, **Figure S1E**) NK cells. Similarly, the frequency of CD8^+^ NK cells were significantly reduced in the patient both at baseline and in response to stimulation (**Figure 3O**, **Figure S1F**), while CD25^+^ NK cells (**Figure 3P**, **Figure S1G**) were normal. This abnormal distribution of markers disproportionately affected CD56^dim^ NK cells more than CD56^bright^ NK cells, although Helios was reduced in both populations (**Figure S1H-N**). Taken together, patient NK cells are skewed towards an immature phenotype.

As NK cells mediate their effector functions through the release of proinflammatory cytokines and cytolysis^33^, we studied whether these roles were affected in P1. In response to stimulation, P1 had significantly elevated IFN-γ^+^ NK cells (**Figure 3Q**, **Figure S1O**), but comparable TNF-α^+^ NK cells (**Figure 3R**, **Figure S1P**). Similarly, P1 NK cells expressed significantly less granzyme B (**Figure 3S**) and perforin (**Figure 3T**), with this effect being more pronounced in CD56^dim^ NK cells (**Figure S1Q-R**). These defects were confirmed in multidimensional space when we clustered live CD3^-^CD56^+^ NK cells from P1 and adult and pediatric controls on CD16, CD27, CD57, CD94, Helios, perforin, granzyme B, IFN-γ, and TNF-α at baseline and in response to stimulation (**Figure 3U-V**, **Figure S1S**).

### Patient CD8^+^ T cells are significantly reduced, have high PD-1 expression, and are potent proinflammatory cytokine producers

As CD8^+^ T cells are frequently enriched in autoimmune disease^36^ and can be clonally expanded in AIHA^37^, we assessed the frequency of CD8^+^ T cells and subsets and their Helios expression. P1 had profound CD8^+^ T cell lymphopenia (**Figure 4A-B**), significantly elevated frequencies of central memory (CM) with a concurrent reduction in naïve subsets, while effector memory (EM) and TEMRA subsets were normal (**Figure 4C-G**). Importantly, all P1 CD8^+^ T cell subsets were Helios-deficient (**Figure 4H-I**).

**Figure 4.**
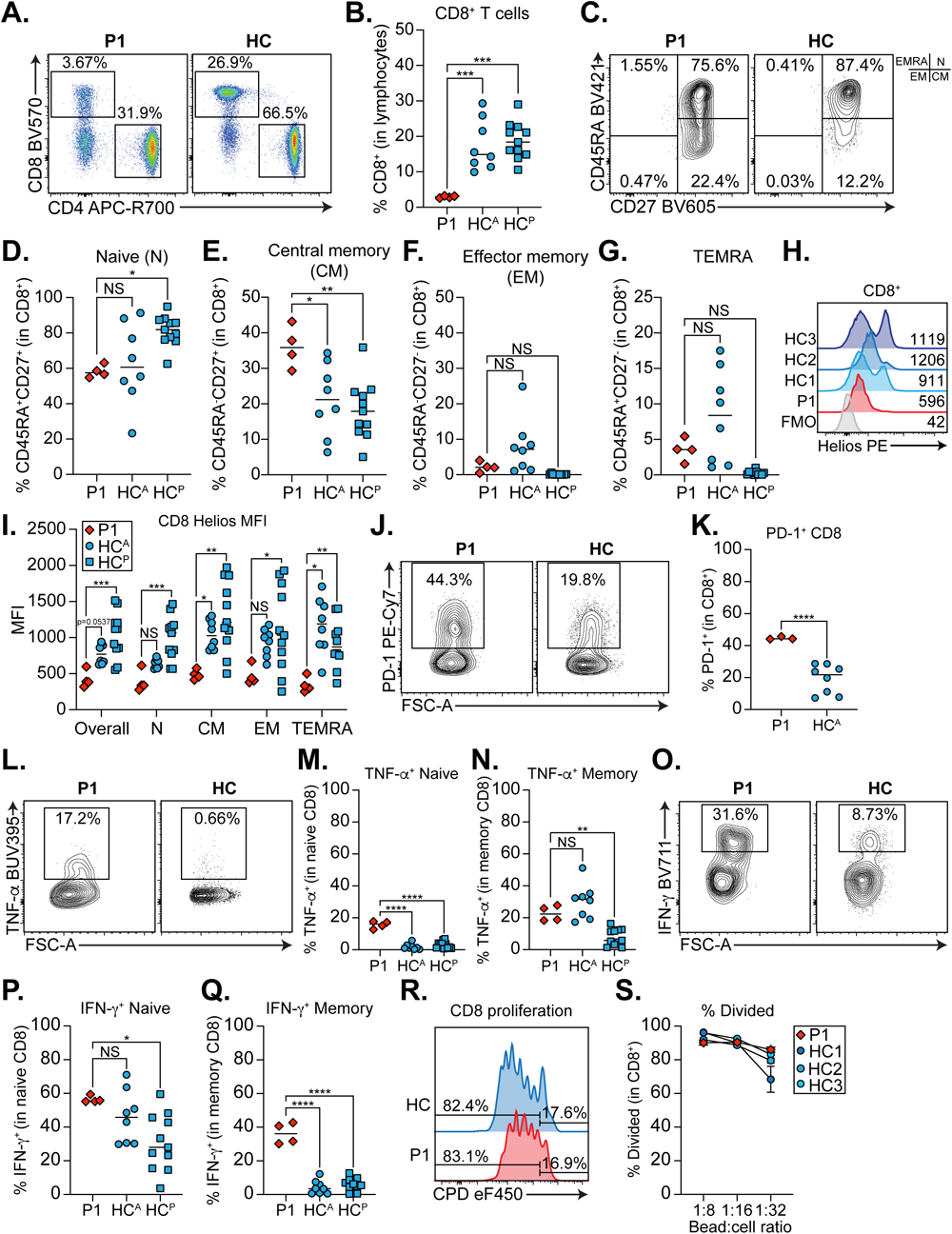
P1 CD8^+^ T cells are more differentiated and have enhanced cytokine production. A) Representative dot plots showing CD3^+^CD4^-^CD8^+^ and CD3^+^CD4^+^CD8^-^ T cells. B) Frequency of CD8^+^ T cells in P1 and adult (HC^A^) or pediatric (HC^P^) controls. C) Representative contour plot of naïve (N), central memory (CM), effector memory (EM), and TEMRA CD8^+^ T cells in the patient and a control. Quadrants corresponding to each subset are shown to the right. D-G) Quantification of D) naive, E) CM, F) EM, G) TEMRA CD8^+^ T cells from C). H) Representative Helios histograms for CD8^+^ T cells from P1 and three controls compared to a fluorescence minus one (FMO) control. Mean fluorescence intensities (MFI) are indicated. I) Quantification of Helios MFI in different CD8^+^ T cell subsets. J) Representative contour plots for PD-1^+^ CD8^+^ T cells in P1 and a control. K) Quantification of J). L-Q) PBMCs stimulated 4h with PMA+ionomycin. L), O) Representative contour plots for L) TNF-α^+^ and O) IFN-γ^+^ CD8^+^ T cells in P1 and a control. M-N) Quantification of TNF-α^+^ M) naïve and N) total memory CD8^+^ T cells. P-Q) Quantification of IFN-γ^+^ P) naïve and Q) total memory CD8^+^ T cells. A-Q) P1 n=5, HC^A^ n=8, HC^P^ n=11. J-K) P1 n=3, HC^A^ n=8. *p<0.05, **p<0.01, ***p<0.001, ****p<0.0001. Ordinary one-way ANOVA with Šidák’s multiple comparisons test. R) Representative histograms for dilution of Cell Proliferation Dye (CPD) eF450 in P1 or control CD8^+^ T cells after 4 days of stimulation with anti-CD3/CD28 beads at 1:32 bead to cell ratio. S) Quantification of percent divided CD8^+^ T cells at 1:8, 1:16, 1:32 bead to cell ratios. Shown are technical duplicates.

PD-1 is a critical inhibitory receptor that is upregulated in activated, memory, or exhausted T cells to restrain effector functions^38^. We therefore assessed PD-1 expression and proinflammatory cytokine production in P1 CD8^+^ T cells. In line with elevated CD8 CM, we discovered significantly more PD-1 expression on P1 CD8^+^ T cells (**Figure 4J-K**). These T cells are likely not exhausted as both naïve and total memory CD8^+^ T cells produced significantly more TNF-α (**Figure 4L-N**) and IFN-γ (**Figure 4O-Q**) than controls, while CD8^+^ T cell proliferation was comparable to controls (**Figure 4R-S**).

### P1 CD19^+^ B cells show largely normal development but increased TNF-**α** production

Although Ikaros and Aiolos are both critical for B cell differentiation^39^, the role of Helios in these cells is less clear. Helios silencing is thought to maintain B cell function as ectopic expression leads to B cell hyperresponsiveness and lymphomagenesis^40^. We therefore studied the development and function of P1 B cells. In general, B cell development was largely normal, with intact total CD19^+^ (**Figure S2A-B**), naïve (**Figure S2C, D**), switched memory [SM] (**Figure S2C, F**), and plasmablasts (**Figure S2I-J**). However, we did observe significantly increased non-switched memory [NSM] (**Figure S2C, E**) and decreased transitional B cells (**Figure S2G-H**), while Helios was lowly expressed in all B cell populations from both P1 and controls (**Figure S2K-L**). In response to stimulation, patient B cells produced significantly more TNF-α than controls (**Figure S2M**). This applied to NSM (**Figure S2N**), SM (**Figure S2O**), and naïve (**Figure S2P**) subsets.

### P1 CD4^+^ T cells are more mature

CD4^+^ T helper (T_H_) cells and their subsets are critical for both protective antimicrobial immunity and driving pathological states such as autoimmunity and atopic disease^41^. We therefore enumerated the frequency of total CD4^+^ T cells and subsets and Helios expression. P1 CD4^+^ T cells were modestly elevated (**Figure 5A**) in line with CD8^+^ T lymphopenia. We also found similar subset distributions to P1 CD8^+^ T cells, including significantly increased CD4^+^ CM (**Figure 5B, D**), with an associated reduction in naïve (**Figure 5B, C**), while EM and TEMRA subsets (**Figure 5E-F**) were normal. Helios expression was low in all CD4^+^ subsets (**Figure 5G-H**).

**Figure 5.**
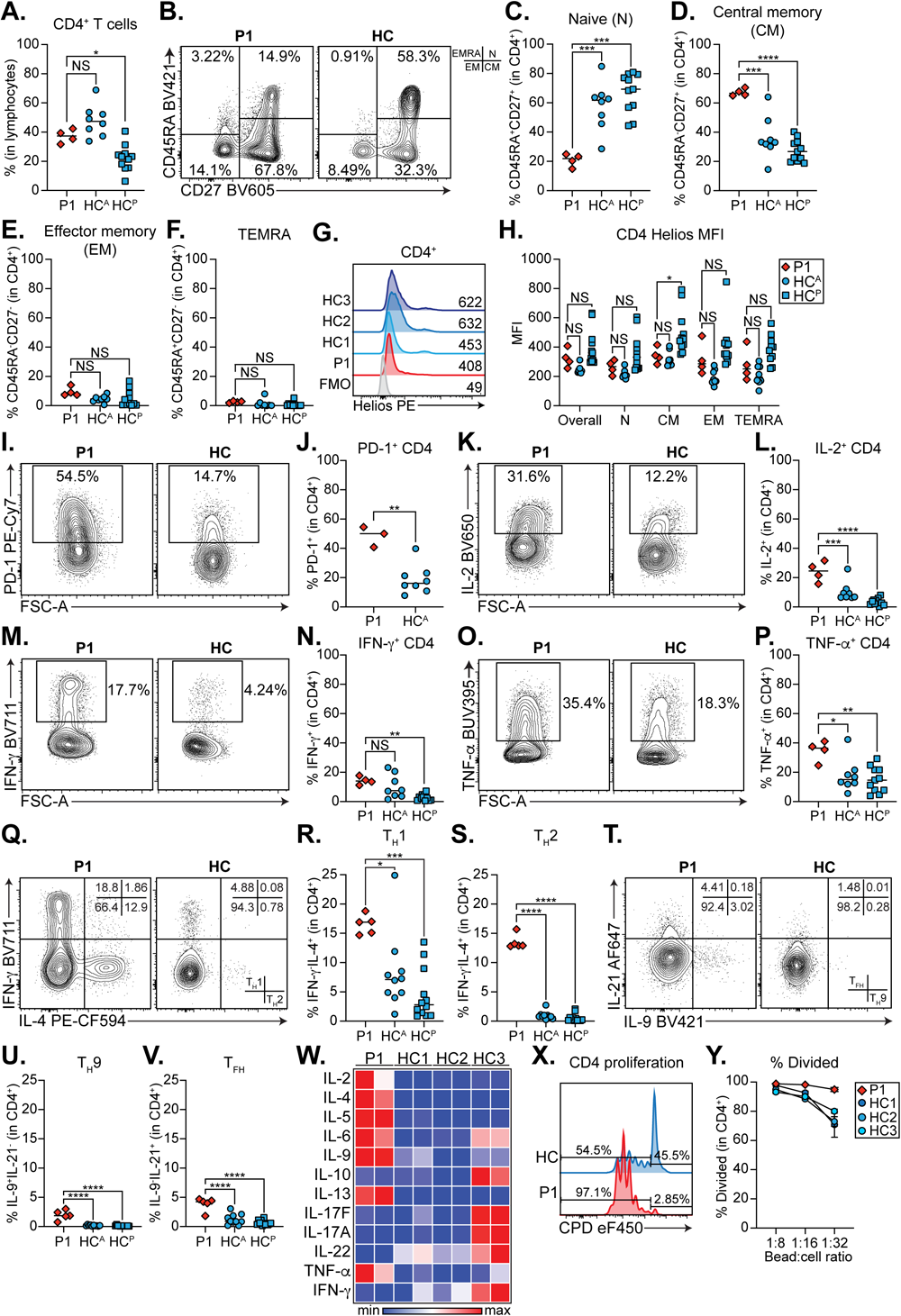
P1 CD4^+^ T cells are hyperactive and exhibit enhanced effector function. _A)_ Quantification of frequency of CD4^+^ T cells in P1 and adult (HC^A^) or pediatric (HC^P^) controls. B) Representative contour plot of naïve (N), central memory (CM), effector memory (EM), and TEMRA CD4^+^ T cells in the patient and a control. Quadrants corresponding to each subset are shown to the right. C-F) Quantification of C) naïve, D) CM, E) EM, F) TEMRA CD4^+^ T cells from B). G) Representative Helios histograms for CD4^+^ T cells in P1 and three controls compared to a fluorescence minus one (FMO) control. Mean fluorescence intensities (MFI) are indicated. H) Quantification of Helios MFI in different CD4^+^ T cell subsets. I) Representative contour plots for PD-1^+^ CD4^+^ T cells in P1 and a control. J) Quantification of I). K-V) PBMCs stimulated 4h with PMA+ionomycin. K), M), O), Q), T) Representative contour plots for K) IL-2^+^ CD4^+^ M) IFN-γ^+^ CD4^+^, O) TNF-α^+^ CD4^+^, Q) IFN-γ^+^IL-4^-^ T_H_1 and IFN-γ^-^IL-4^+^ T_H_2, T) IL-21^+^IL-9^-^ T_FH_ and IL-21^-^IL-9^+^ T_H_9 cells in P1 and a control. Quadrant identities are shown on the bottom right. Frequencies corresponding to each quadrant are included to the top right of each panel. L), N), P), R-S), U-V) Quantification of L) IL-2^+^ CD4^+^, N) IFN-γ^+^ CD4^+^, P) TNF-α^+^ CD4^+^, R) T_H_1, S) T_H_2), U) T_H_9, V) T_FH_ cells. A-H), K-V) P1 n=5, HC^A^ n=8, HC^P^ n=11. I-J) P1 n=3, HC^A^ n=8. *p<0.05, **p<0.01, ***p<0.001, ****p<0.0001. Ordinary one-way ANOVA with Šidák’s multiple comparisons test. W-Y) PBMCs stimulated with anti-CD3/CD28 beads at various bead to cell ratios for 4 days. W) T_H_ cytokines measured by LEGENDplex. X) Representative histograms for dilution of Cell Proliferation Dye (CPD) eF450 in P1 or control CD4^+^ T cells stimulated at 1:32 bead to cell ratio. Y) Quantification of percent divided CD4^+^ T cells at 1:8, 1:16, 1:32 bead to cell ratios. Shown are technical duplicates.

### P1 CD4^+^ T cells are hyperresponsive and exhibit effector phenotypes

In mice, Treg-specific, but not complete knockout, of *Ikzf2* in mice leads to increased PD-1 expression on CD4^+^ T cells^20^. Surprisingly, P1 had three-fold higher PD-1^+^ CD4^+^ T cells than controls (**Figure 5I-J**), which could signify higher activation status. We thus measured the frequency of T_H_ subsets and cytokine production. Mirroring our *IL2* promoter luciferase data (**Figure 2F-G**), we observed a striking increase in IL-2^+^ CD4^+^ T cells in P1 (**Figure 5K-L**). This was not limited to IL-2, as the patient also had significantly increased IFN-γ^+^ and TNF-α^+^ CD4^+^ T cells (**Figure 5M-P**) and most T_H_ subsets, including T_H_1, T_H_2 (**Figure 5Q-S**), T_H_9, T_FH_ (**Figure 5T-V**), and T_H_17 (**Figure S3A-B**). This also held true when we measured T_H_ cytokines in the supernatant of stimulated PBMCs, where we found markedly elevated concentrations of IL-2, IL-4, IL-5, IL-6, IL-9, IL-13, and TNF-α, even in response to lower bead:cell ratios (**Figure 5W, Figure S3C-N**). Surprisingly, we observed marked impairment in secreted IL-22 levels (**Figure S3L**). In keeping with this picture of hyperactive CD4^+^ T cells, P1 CD4^+^ T cells more readily proliferated in response to stimulation than controls (97.1% divided in patient vs. 54.5% in control) (**Figure 5X-Y**).

### P1 has normal Treg frequencies and TSDR methylation

∼70-80% of Tregs express Helios and at least in mice, this is thought to be important for regulating Treg identity and function^42^. As Tregs are critical mediators of tolerance/homeostasis and their dysfunction is associated with allergic and autoimmune disease^43^, we enumerated their frequency and Helios expression in P1. P1 had normal frequencies of CD3^+^CD4^+^CD8^-^CD25^+^CD127^lo^FOXP3^+^ Tregs (**Figure 6A, Figure S4A**), but significantly reduced Helios expression (**Figure 6B-C**).

**Figure 6.**
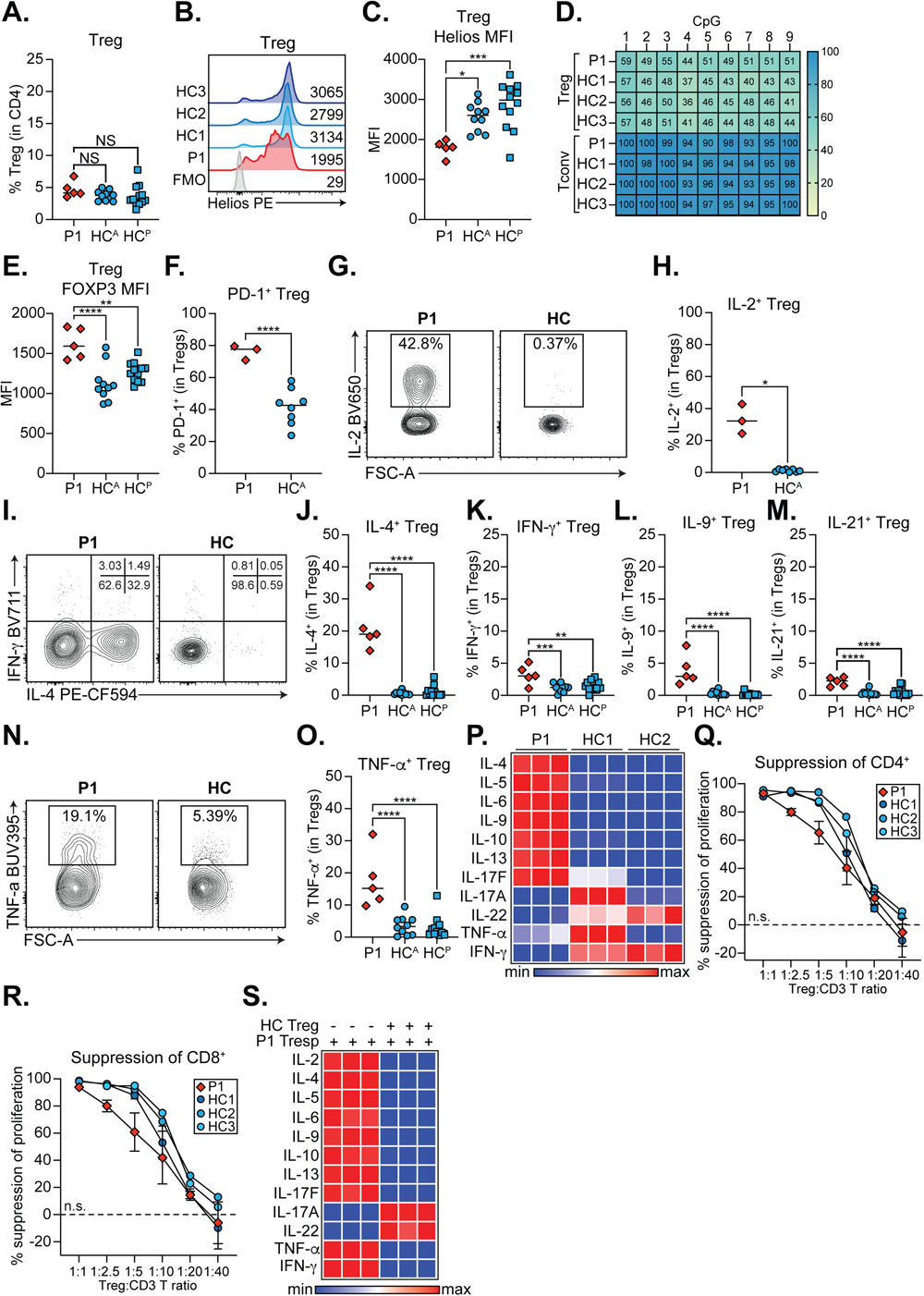
P1 Tregs have aberrant cytokine production and impaired suppressive function. A) Quantification of frequency of CD3^+^CD4^+^CD8^-^CD25^+^CD127^lo^FOXP3^+^ Tregs in P1 and adult (HC^A^) or pediatric (HC^P^) controls. B) Representative Helios histograms for Tregs in P1 and three controls compared to a fluorescence minus one (FMO) control. Mean fluorescence intensities (MFI) are indicated. C) Quantification of Helios MFI in Tregs. D) CpG methylation of the Treg-specific demethylated region (TSDR) for P1 or control Tregs and Tconvs. Percent methylation at different TSDR CpG sites is indicated. Representative of n=2. E) Quantification of FOXP3 MFI in Tregs. F) Quantification of PD-1^+^ Tregs. G-O) PBMCs stimulated 4h with PMA+ionomycin. G), I), N) Representative contour plots for G) IL-2^+^, I) IFN-γ^+^, IL-4^+^, and N) TNF-α^+^ Tregs. H), J-M), O) Quantification of H) IL-2^+^, J) IL-4^+^, K) IFN-γ^+^, L) IL-9^+^, M) IL-21^+^, O) TNF-α^+^ Tregs. A-C), E), J-O) P1 n=5, HC^A^ n= 10, HC^P^ n=12. F-H) P1 n=3, HC^A^ n=8. *p<0.05, **p<0.01, ***p<0.001, ****p<0.0001. Ordinary one-way ANOVA with Šidák’s multiple comparisons test. P) Isolated and expanded Tregs stimulated with anti-CD3/CD28 beads for 4 days. T_H_ cytokines measured by LEGENDplex. Shown are 3 technical replicates. Q-R) Isolated and expanded Tregs co-cultured at different ratios with control CD3^+^ T responder (Tresp) cells. n=2. Q-R) Suppression of Q) CD4^+^ and R) CD8^+^ T cell proliferation. Significance determined by one-way ANOVA of areas under the curve. S) T_H_ cytokines measured by LEGENDplex when co-culturing 1:16 bead:cell ratio stimulated P1 CD3^+^ Tresp with control Tregs. Shown are technical triplicates.

Since methylation of the Treg-specific demethylated region (TSDR) in the *FOXP3* gene is important for maintaining stable FOXP3 expression and Treg suppressive function^44,45^, we quantified expanded P1 Treg TSDR methylation and FOXP3 expression in comparison to Tregs from healthy female controls. TSDR methylation was normal (**Figure 6D**), although the average methylation trended to higher in P1 (**Figure S4B**). E*x vivo* P1 Tregs had significantly higher FOXP3 expression than controls, likely reflecting a higher state of activation (**Figure 6E**).

### P1 Tregs are predominantly PD-1^+^ and are potent T_H_ cytokine producers

Given the striking increase in PD-1^+^ CD4+ (**Figure 5I-J**) and CD8^+^ (**Figure 4J-K**) T cells in P1, we assessed PD-1 expression in P1 *ex vivo* Tregs. Similarly, P1 had significantly more PD-1^+^ Tregs (∼80%) than controls (∼40%) (**Figure 6F, Figure S4C**). Helios genetic or pharmacological targeting in Tregs leads to elevated IL-2^32^, IFN-γ, and IL-17A^19,20,28,46^ and effector T_H_ cell signatures^47^. We thus studied T_H_ cytokine production in *ex vivo* Tregs and discovered significantly elevated IL-2^+^ Tregs in P1 **(Figure 6G-H)**, confirming our *IL2* luciferase data (**Figure 2F-G**). P1 also had a striking increase in IL-4^+^ (**Figure 6I-J**), IFN-γ^+^ (**Figure 6I, K**), IL-9^+^ (**Figure 6L, Figure S4D**), IL-21^+^ **(Figure 6M, Figure S4D**), and TNF-α^+^ (**Figure 6N-O**) Tregs. Expanded P1 Tregs also produced higher concentrations of IL-4, IL-5, IL-6, IL-9, IL-10, IL-13, and IL-17F (**Figure 6P, Figure S4C-N**).

### P1 Tregs have impaired suppressive function are not intrinsically resistant to being suppressed

The abnormal cytokine production in P1 Tregs prompted us to investigate their ability to suppress T responder (Tresp) proliferation and cytokine production. We cocultured expanded P1 and control Tregs at different ratios with stimulated control CD3^+^ Tresp. We discovered no significant differences between patient or control Treg-mediated suppression of CD4^+^ (**Figure 6Q**) or CD8^+^ (**Figure 6R**) T cell proliferation, although suppression trended to lower in the 1:2.5 and 1:5 Treg:CD3 T cell ratios.

An intrinsic resistance to Treg-mediated suppression could also contribute to the patient’s prominent hyperactivation phenotype. We reversed our setup from above and cocultured control Tregs with P1 CD3^+^ Tresp and measured suppression of T_H_ cytokines. P1 T_H_ cytokine production was effectively suppressed by control Tregs (**Figure 6S, Figure S5**), excluding an intrinsic defect in susceptibility to suppression.

### scRNA-seq reveals a predominantly memory and effector phenotype in P1 Tregs

To assess transcriptomic pathways that could drive the striking patient Treg phenotype, we sorted CD4^+^CD25^+^CD127^lo^ Tregs from P1 and two age-matched/sex-matched controls (**Figure S6A**). We carried out whole transcriptome single-cell RNA sequencing (scRNA-seq) on Tregs from P1 (n=295 cells) and controls (n=331 cells) and performed dimensionality reduction using the uniform manifold approximation and projection (UMAP) method^48^. Patient Tregs clustered separately from controls (**Figure 7A**) and when cell identity was annotated using the DICE project^49^, they were predominantly labelled as memory Treg and T_H_ subsets (T_H_1 T_H_2, T_H_17, T_FH_), in line with the activated and aberrant cytokine phenotype observed (**Figure 6F-P**). Similarly, annotation based on CD45RA AbSeq/HLA-DR AbSeq expression to identify CD45RA^+^HLA-DR^-^ naïve, CD45RA^-^HLA-DR^+^ activated, and CD45RA^-^HLA-DR^-^ Tregs^50,51^ also found that P1 Tregs were predominantly CD45RA^-^HLA-DR^+^ and CD45RA^-^HLA-DR^-^. (**Figure 7B**). Nevertheless, P1 Treg subsets were transcriptomically similar to control Tregs (**Figure 7C-E**, **Figure S6B-D**), except for significantly increased *IL2*, *PDCD1*, and *CCR4* transcript abundance in P1 naïve Tregs (**Figure 7C**), consistent with our flow cytometry data (**Figure 6F-H**).

**Figure 7.**
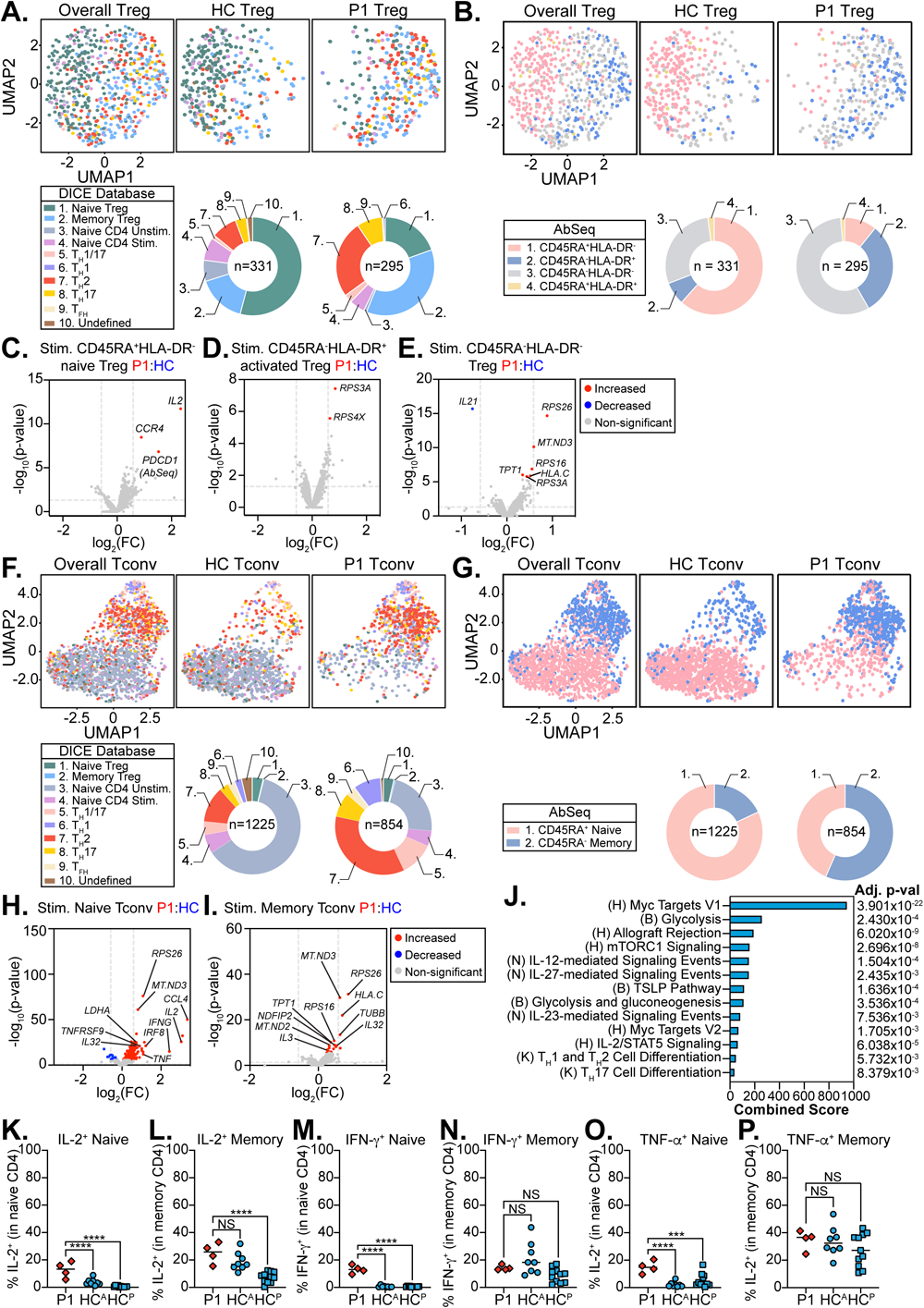
P1 naïve CD4^+^ T cells are more active and primed for effector function. A-E) Single-cell RNA sequencing (scRNA-seq) of sorted CD3^+^CD4^+^CD25^+^CD127^lo^ Tregs from P1 and 2 age-matched and sex-matched controls. A) Unstimulated Tregs clustered and annotated using the DICE project and B) CD45RA/HLA-DR AbSeq labelling. Panels represent the controls and P1 Tregs clustered together (overall) or control or P1 Tregs alone. Included are legends with colours and numbers corresponding to each cell type labelled. Doughnut plots are included below each panel to represent the frequency of each annotated cell type. C-E) Volcano plots comparing stimulated patient and control C) CD45RA^+^HLA-DR^-^ naïve, D) CD45RA^-^HLA-DR^+^ activated, and E) CD45RA^-^HLA-DR^-^ Tregs. Red=significantly increased, blue=significantly decreased, gray=non-significant. Vertical dashed line: fold change=1.5, horizontal dashed line: FDR=0.05. Top 10 most significant genes are labelled. F-H) scRNA-seq of sorted CD3^+^CD4^+^CD25^-^CD127^hi^ Tconv from P1 and 2 age-matched and sex-matched controls. F-G) Unstimulated Tconv clustered and annotated as in A-B). H-I) Volcano plots as in C-E) comparing stimulated patient and control H) CD45RA^+^ naïve or I) CD45RA^-^ memory Tconvs. J) Gene set enrichment of differentially expressed genes between stimulated naïve patient and control Tconvs using EnrichR. Shown are the combined scores and adjusted p-values. H=MSigDB Hallmark, B=BioPlanet 2019, N=NCI Nature 2016, K=KEGG 2021 Human. K-P) PBMCs stimulated 4h with PMA+ionomycin. Quantification of stimulated K) IL-2^+^ naïve, L) IL-2^+^ total memory, M) IFN-γ^+^ naïve, N) IFN-γ^+^ total memory, O) TNF-α^+^ naïve, and P) TNF-α^+^ total memory CD4^+^ T cells. K-P) P1 n=5, HC^A^ n=8, HC^P^ n=11. ***p<0.001, ****p<0.0001. Ordinary one-way ANOVA with Šidák’s multiple comparisons test.

### scRNA-seq reveals a predominantly memory and effector phenotype in P1 CD4^+^ Tconvs

Given the profound CD4 phenotype, we also sorted CD4^+^CD25^-^CD127^+^ Tconv cells from P1 and two age-matched/sex-matched controls and performed scRNA-seq (P1 n=854 cells; control n=1225 cells) (**Figure S7A**). Like Tregs, P1 Tconv clustered separately from controls and were frequently labelled as T_H_ subsets (**Figure 7F**) and CD45RA^-^ memory (**Figure 7G**). When comparing the transcriptome of AbSeq-inferred naïve (**Figure 7H, Figure S7E**) and memory (**Figure 7I, Figure S7F**) CD4^+^ Tconv cells between P1 and controls, we found similar memory, but distinctly different naïve populations. Unstimulated naïve P1 CD4^+^ Tconv cells had significantly higher transcript abundance of genes related to IL-2 signalling, activation, and migration (*IL2RB*, *CXCR4*, *PDCD1*, *CCL5*, *JUN*). Stimulated P1 CD4^+^ Tconv cells had significantly higher expression of proinflammatory cytokine/chemokine genes (*IL2*, *IFNG*, *TNF*, *CCL4*), confirming our CD4^+^ phenotyping data (**Figure 5K-P**).

### P1 CD4^+^ Tconvs are poised for T_H_ differentiation

To study what pathways could be driving naïve CD4^+^ Tconv differences, we carried out gene set enrichment analyses. Unstimulated naïve P1 CD4^+^ Tconv cells were more enriched in pathways related to proliferation, IL-6 signalling, allograft rejection, apoptosis, and hypoxia. Further, stimulated patient CD4^+^ Tconv cells showed significant enrichment in proliferation (Myc targets V1/V2), metabolism (glycolysis, gluconeogenesis, mTORC1 signaling), Treg survival (IL-2-STAT5), impaired tolerance (allograft rejection), atopy (TSLP pathway), and T_H_ differentiation (IL-27-/IL-12-/IL-23-mediated signalling events, T_H_1, T_H_2, T_H_17 cell differentiation) pathways (**Figure 7J**). In line with the primed/poised phenotype of P1 naïve CD4^+^ Tconv cells, we observed significantly increased IL-2^+^ (**Figure 7K-L**), IFN-γ^+^ (**Figure 7M-N**), and TNF-α^+^ (**Figure 7O-P**) naïve P1 CD4^+^ T cells, with a similar trend observed in memory.

## DISCUSSION

Germline pathogenic variants in *IKZF2* have only been discovered in humans in the past two years. The initial descriptions were of germline heterozygous or homozygous LOF *IKZF2* variants in a total of 13 patients with combined immunodeficiency and/or immune dysregulation^7–9^ (summarized in **Figure S7A, Table S3**). Collectively, these germline LOF *IKZF2* variants were predominantly heterozygous (10/13) and were mostly associated with evidence of immune dysregulation (10/13) (**Table S4**), including systemic lupus erythematosus (SLE), hemophagocytic lymphohistiocytosis (HLH), idiopathic thrombocytopenic purpura (ITP), and Evan’s syndrome. Our group recently expanded this list through the discovery of 2 patients with germline DN *IKZF2* variants who presented with ICHAD syndrome^10^. Notably the patient carrying the DN pathogenic c.406+540_574+13477dup;p.Gly136_Ser191dup *IKZF2* variant (designated P1) experienced the most severe clinical manifestations of immune dysregulation out of all 15 patients described to date with pathogenic germline *IKZF2* variants (**Figure S7B**). P1 presented with syndromic developmental features plus significant chronic AIHA and atopic dermatitis, features consistent with classification as a PIRD. Given the profound immune dysregulation associated with the DN pathogenic c.406+540_574+13477dup;p.Gly136_Ser191dup *IKZF2* variant, we embarked on a detailed immunological assessment of P1.

Immunophenotyping of this patient with DN *IKZF2* deficiency revealed striking differences from previous homozygous and heterozygous LOF patients and mice. For example, P1 NK cells were phenotypically immature and likely hyperactive due to elevated IFN-γ production and reduced intracellular perforin and granzyme B expression (**Figure 3**). In contrast, other reported patients had NK cell lymphopenia^8,9^ or normal NK development^7^. Furthermore, while *IKZF2*^-/-^ mice have normal NK cell frequencies^21^, NK cells from NKp46-mutant mice (Noé) show elevated Helios expression and effector function, which modulates protective memory CD4^+^/CD8^+^ T cell development^52^. The study of P1 has thus revealed a previously unappreciated role for Helios in human NK cell development and function and calls for more in-depth studies in the future.

Helios has been extensively studied in mouse Tregs^42^, but its role in human Tregs has remained relatively unclear. Previous CRISPR-mediated KO studies by our co-authors found that Helios is dispensable for lineage stability and suppressive function in fully differentiated Tregs^27^, whereas germline homozygous or heterozygous LOF *IKZF2* variants caused a proinflammatory Treg phenotype associated with increased IL-2 and IFN-γ production, but intact suppressive function^7,8^. Our functional studies with this DN *IKZF2* variant confirm that it is not required for Treg development, but that it is essential for inhibition of IL-2 expression and many other cytokines. Although functional suppression of T cell proliferation appeared normal, aberrant production of many T_H_ cytokines likely contributes to a functional defect in immune regulation *in vivo* (**Figure 6**), which is exacerbated by the greater propensity of P1 naïve CD4^+^ T cells to differentiate into effector CD4^+^ T cell subsets (T_H_1, T_H_2, T_H_9, T_FH_).

Taken together, we have defined the immune phenotype of a novel form of Helios deficiency caused by a DN variant, which leads to severe congenital AIHA and atopic dermatitis^10^. This study identifies new players in the pathogenesis of these immune-mediated disorders and has significantly expanded our understanding of human Helios, including its regulation of naïve CD4 T cell differentiation into memory/effector subsets and NK cell development and function.

## Data Availability

All data produced in the present study are available upon reasonable request to the authors.

## ACKNOWLEDGEMENTS

This work was supported in part by grants from the Canadian Institutes of Health Research (PJT 178054 to S.E.T.; FDN-154304 to M.K.L.), Genome British Columbia (SIP007) (S.E.T.), and BC Children’s Hospital Foundation. S.E.T. holds a Tier 1 Canada Research Chair in Pediatric Precision Health and the Aubrey J. Tingle Professor of Pediatric Immunology. M.K.L. receives a BCCHR salary award and holds a Tier 1 Canada Research Chair in Engineered Immune Tolerance. H.Y.L. is supported by a CIHR Frederick Banting and Charles Best Canada Graduate Scholarships Doctoral Award (CGS-D), University of British Columbia Four Year Doctoral Fellowship (4YF), Killam Doctoral Scholarship, Friedman Award for Scholars in Health, and a BC Children’s Hospital Research Institute Graduate Studentship. M.V.S. is supported by a Vanier Canada Graduate Scholarship and 4YF. A.J.L. is supported by a CGS-D and 4YF. M.S. is supported by a CGS-D and 4YF. We would like to acknowledge the Flow Cytometry and Imaging cores at BC Children’s Hospital Research Institute for providing the equipment for microscopy, flow cytometry, and cell sorting, the Biomedical Research Centre Sequencing Core (BRC-Seq) for their assistance with single-cell RNA sequencing and data processing, The Centre for Applied Genomics, The Hospital for Sick Children, Toronto, Canada for their assistance with whole genome sequencing, and the BC Children’s Hospital BioBank for providing access to age-matched and sex-matched control peripheral blood mononuclear cells.

## AUTHORSHIP CONTRIBUTIONS

H.Y.L, A.L., and S.E.T. were responsible for study conception and design. H.Y.L., M.V.S., A.J.L., M.S., J.G., G.X.Y., S.L. performed experiments. A.M., C.V., A.L., S.M. conducted genetic screening and variant prioritization. H.Y.L., M.S., M.P.F., M.S.K., A.M performed bioinformatics analyses or provided computational resources. H.Y.L., M.V.S., A.S., R.R., L.A., J.H., C.L.Y., M.C., E.H., Au.S., F.K.K., K.J.H., C.M.B., C.V., A.L., S.E.T. collected and analyzed clinical data. H.Y.L., M.V.S., A.J.L., M.S., A.M., J.G., G.X.Y., M.K.L., S.E.T. analyzed and interpreted data. H.Y.L. and S.E.T. wrote the manuscript with input from all authors. All authors read and approved the final version of the manuscript.

---

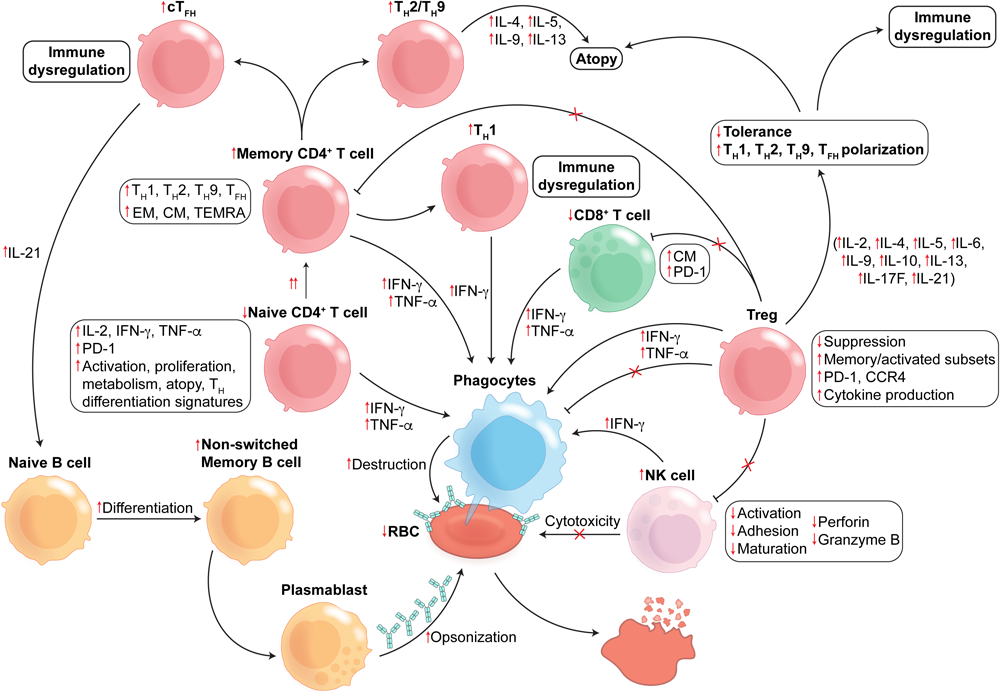

