## Supplemental_Tables_and_Figures for "A germline heterozygous dominant negative *IKZF2* variant causing syndromic primary immune regulatory disorder and ICHAD"

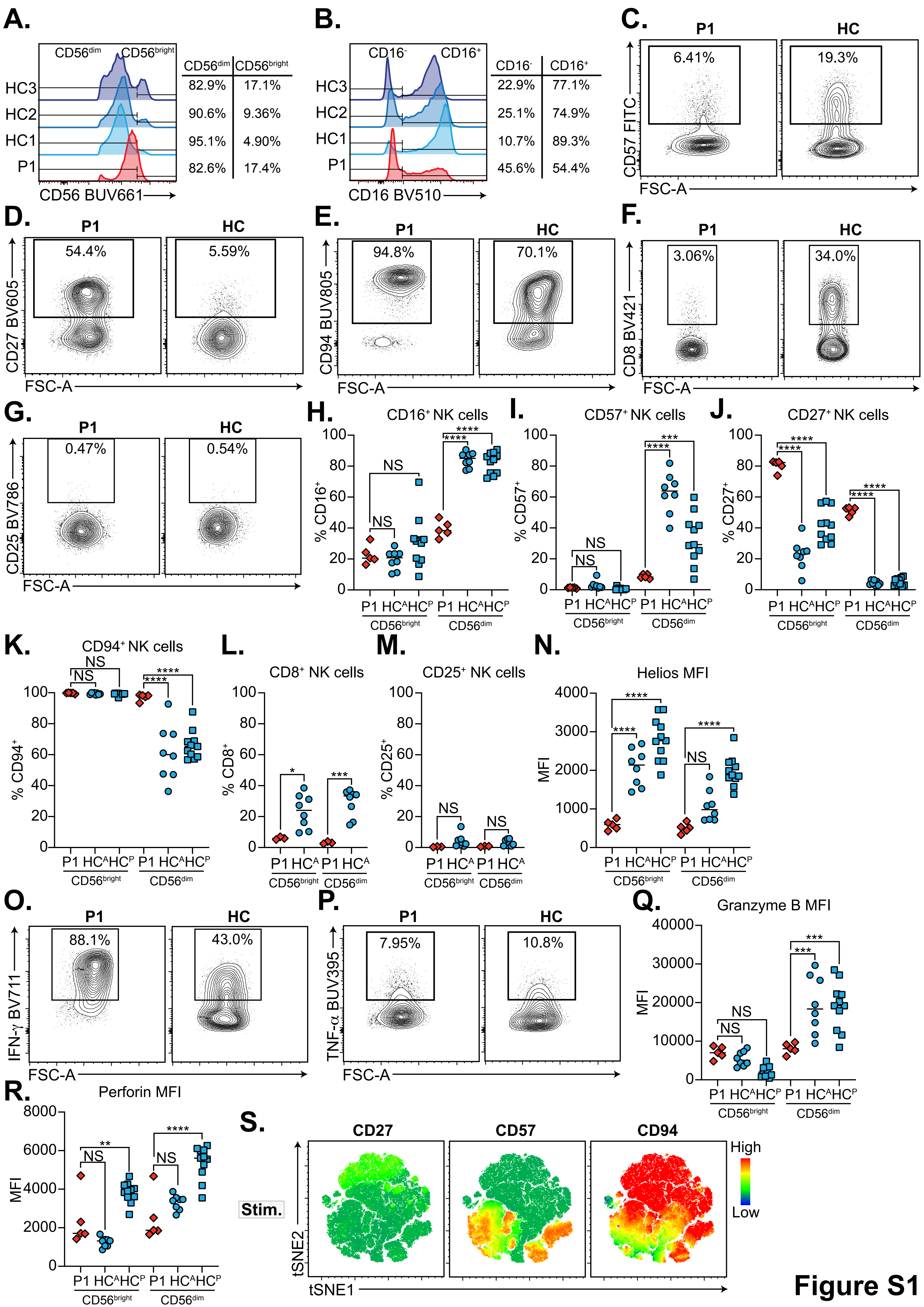
A.
CD56dim CD56bright
HC3 82.9% 17.1%
HC2 90.6% 9.36%
HC1 95.1% 4.90%
P1 82.6% 17.4%
CD56 BUV661
B.
CD16- CD16+
HC3 22.9% 77.1%
HC2 25.1% 74.9%
HC1 10.7% 89.3%
P1 45.6% 54.4%
CD16 BV510
C.
P1 6.41% HC 19.3%
CD57 FITC
FSC-A
D.
P1 54.4% HC 5.59%
CD27 BV605
FSC-A
E.
P1 94.8% HC 70.1%
CD94 BUV805
FSC-A
F.
P1 3.06% HC 34.0%
CD8 BV421
FSC-A
G.
P1 0.47% HC 0.54%
CD25 BV786
FSC-A
H.
CD16+ NK cells
% CD16+
I.
CD57+ NK cells
% CD57+
J.
CD27+ NK cells
% CD27+
K.
CD94+ NK cells
% CD94+
L.
CD8+ NK cells
% CD8+
M.
CD25+ NK cells
% CD25+
N.
Helios MFI
MFI
O.
P1 88.1% HC 43.0%
IFN-γ BV711
FSC-A
P.
P1 7.95% HC 10.8%
TNF-α BUV395
FSC-A
Q.
Granzyme B MFI
MFI
R.
Perforin MFI
MFI
P1 HCA HCP
CD56bright CD56dim
S.
CD27 CD57 CD94
Stim.
High
Low
tSNE2
tSNE1
Figure S1

### Figure S2

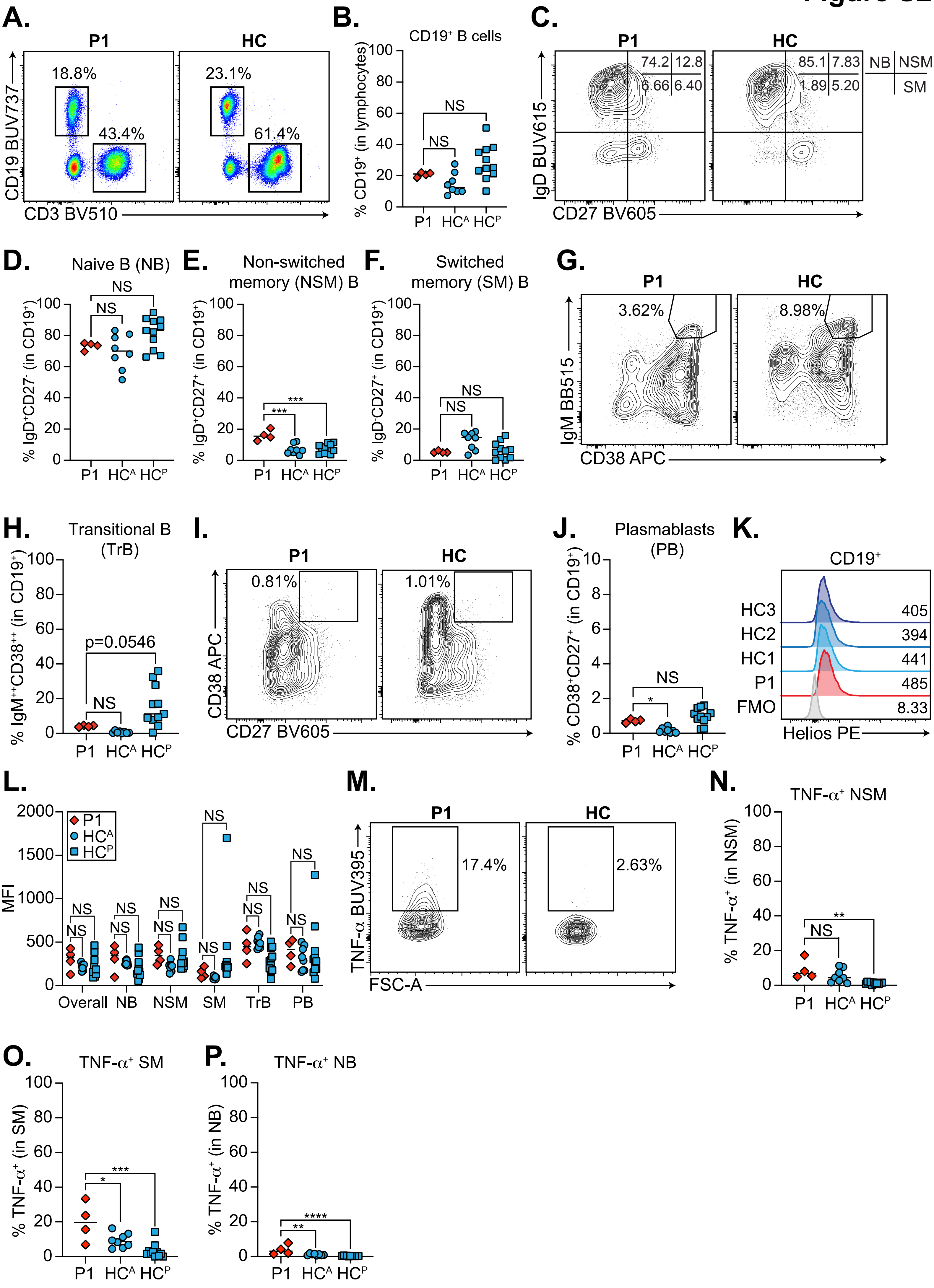

A.
P1
HC
18.8%
43.4%
23.1%
61.4%
CD19 BUV737
CD3 BV510
B.
CD19+ B cells
% CD19+ (in lymphocytes)
NS
NS
P1 HCA HCP
C.
P1
HC
74.2 12.8
6.66 6.40
85.1 7.83
1.89 5.20
NB NSM
SM
IgD BUV615
CD27 BV605
D.
Naive B (NB)
% IgD+CD27- (in CD19+)
E.
Non-switched memory (NSM) B
% IgD+CD27+ (in CD19+)
F.
Switched memory (SM) B
% IgD-CD27+ (in CD19+)
G.
3.62%
8.98%
IgM BB515
CD38 APC
H.
Transitional B (TrB)
% IgM++CD38++ (in CD19+)
p=0.0546
I.
0.81%
1.01%
CD38 APC
CD27 BV605
J.
Plasmablasts (PB)
% CD38+CD27+ (in CD19+)
K.
CD19+
HC3 405
HC2 394
HC1 441
P1 485
FMO 8.33
Helios PE
L.
MFI
P1
HCA
HCP
Overall NB NSM SM TrB PB
M.
17.4%
2.63%
TNF-α BUV395
FSC-A
N.
TNF-α+ NSM
% TNF-α+ (in NSM)
O.
TNF-α+ SM
% TNF-α+ (in SM)
P.
TNF-α+ NB
% TNF-α+ (in NB)

### Figure S3

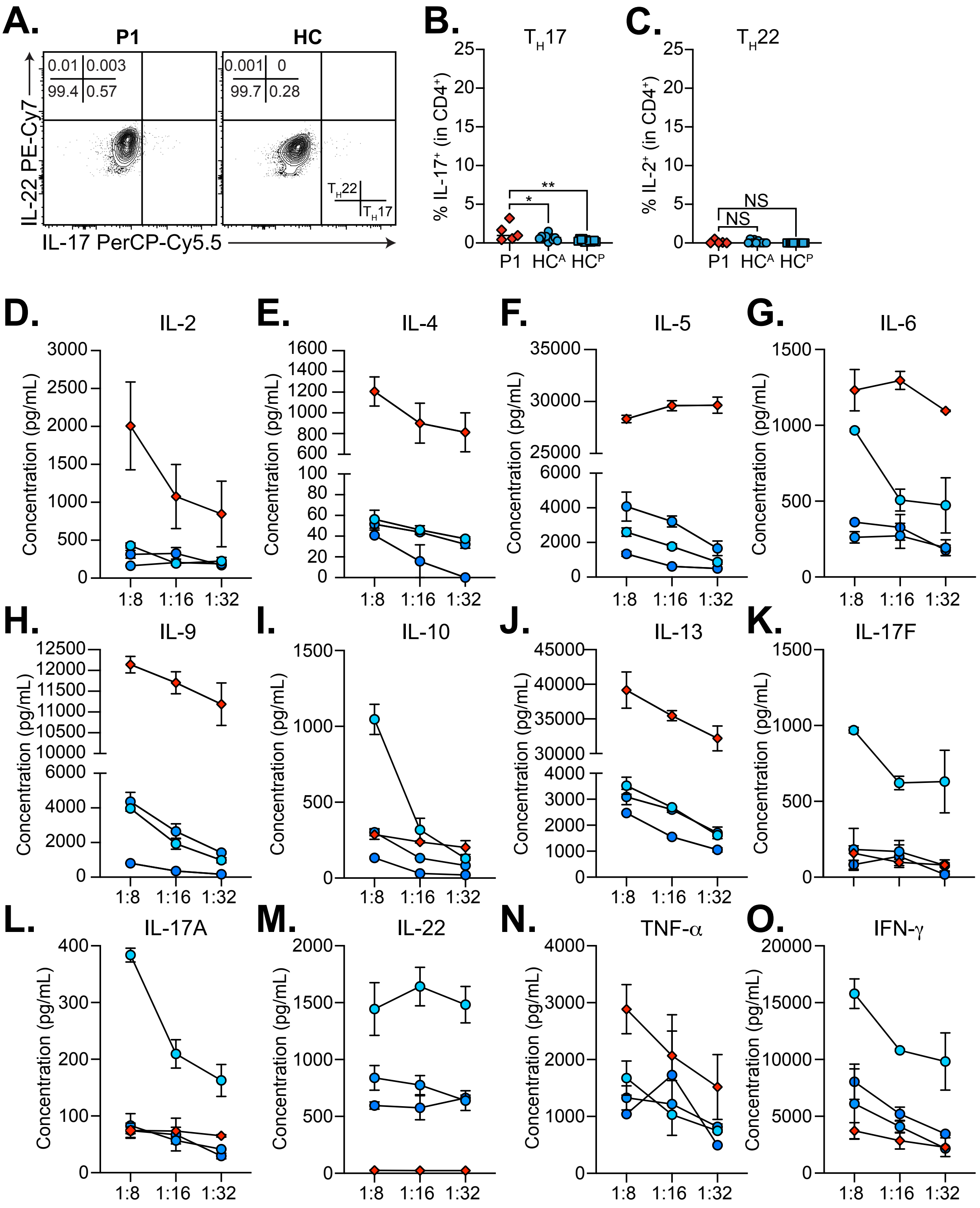
A.
P1
HC
0.01
0.003
99.4
0.57
0.001
0
99.7
0.28
TH22
TH17
IL-22 PE-Cy7
IL-17 PerCP-Cy5.5
B.
TH17
% IL-17+ (in CD4+)
P1
HCA
HCP
*
**
C.
TH22
% IL-2+ (in CD4+)
NS
NS
D.
IL-2
E.
IL-4
F.
IL-5
G.
IL-6
H.
IL-9
I.
IL-10
J.
IL-13
K.
IL-17F
L.
IL-17A
M.
IL-22
N.
TNF-α
O.
IFN-γ
Concentration (pg/mL)
1:8
1:16
1:32

### Figure S4

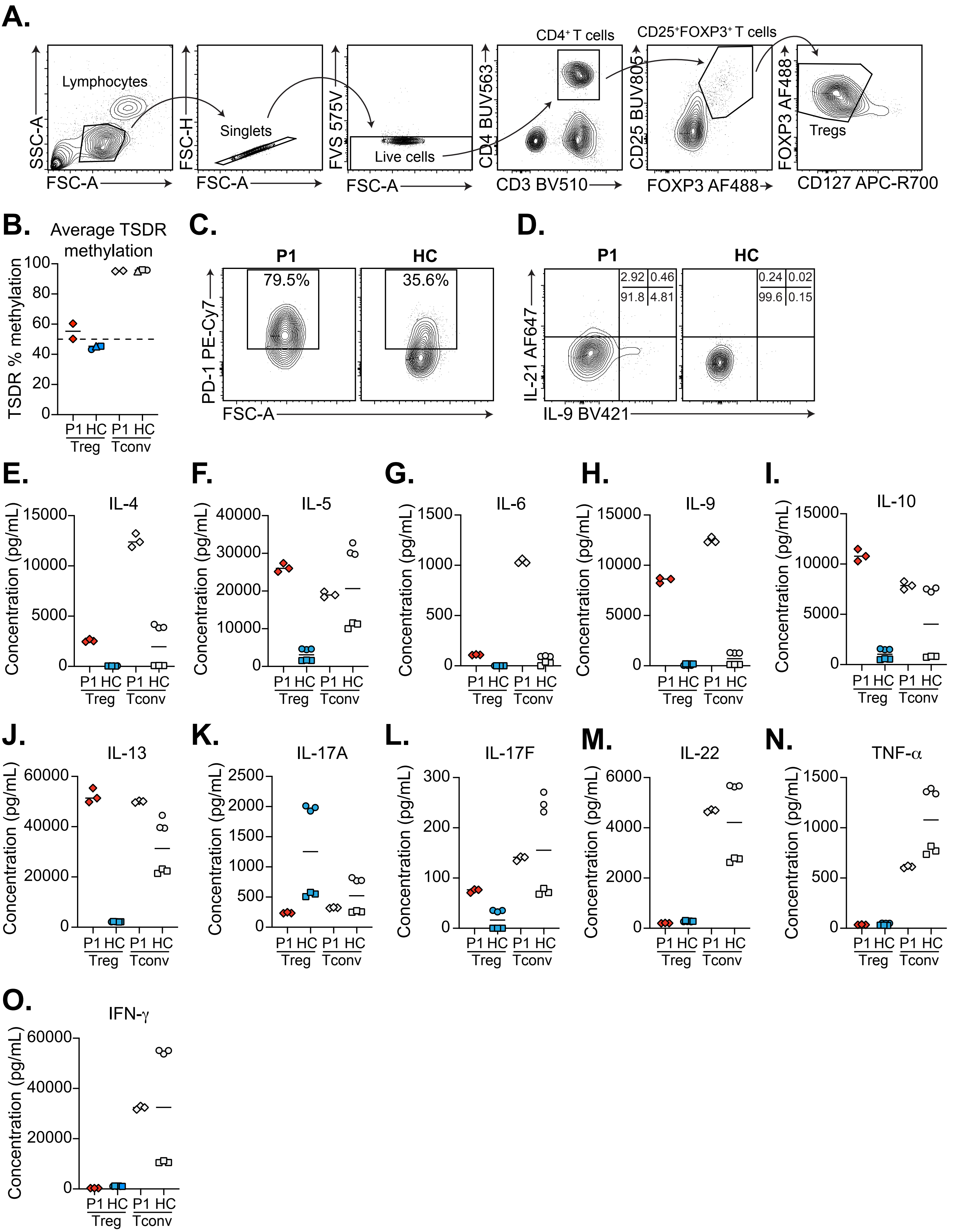
A.
Lymphocytes
SSC-A
FSC-A
Singlets
FSC-H
FSC-A
FVS 575V
Live cells
FSC-A
CD4⁺ T cells
CD4 BUV563
CD3 BV510
CD25⁺FOXP3⁺ T cells
CD25 BUV805
FOXP3 AF488
FOXP3 AF488
Tregs
CD127 APC-R700
B.
Average TSDR methylation
TSDR % methylation
P1 HC P1 HC
Treg Tconv
C.
P1
HC
79.5%
35.6%
PD-1 PE-Cy7
FSC-A
D.
P1
HC
2.92 0.46
91.8 4.81
0.24 0.02
99.6 0.15
IL-21 AF647
IL-9 BV421
E. IL-4
F. IL-5
G. IL-6
H. IL-9
I. IL-10
J. IL-13
K. IL-17A
L. IL-17F
M. IL-22
N. TNF-α
O. IFN-γ
Concentration (pg/mL)
P1 HC P1 HC
Treg Tconv

### Figure S5

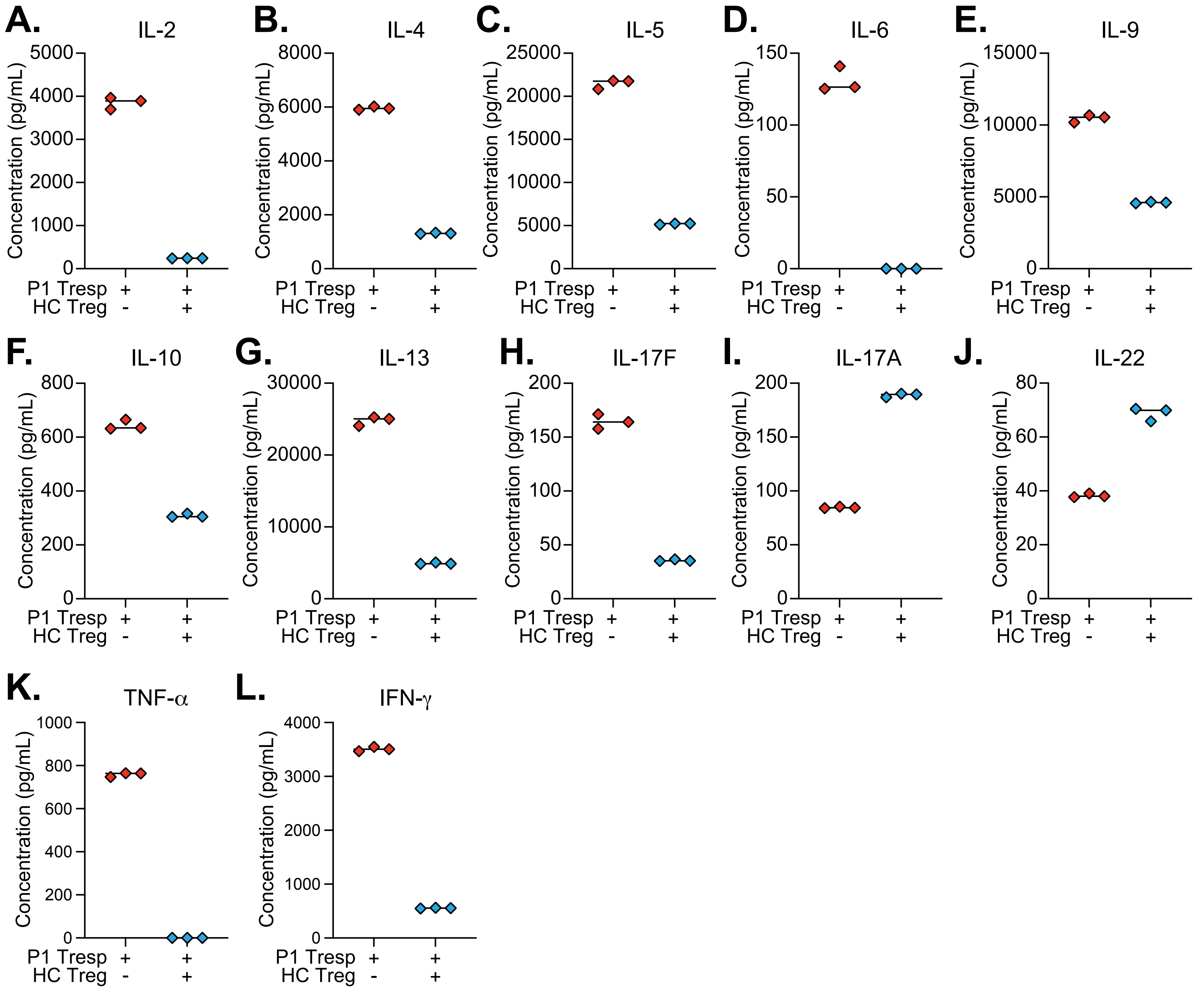

A.
IL-2
B.
IL-4
C.
IL-5
D.
IL-6
E.
IL-9
F.
IL-10
G.
IL-13
H.
IL-17F
I.
IL-17A
J.
IL-22
K.
TNF-α
L.
IFN-γ
Concentration (pg/mL)
P1 Tresp
HC Treg

### Figure S6

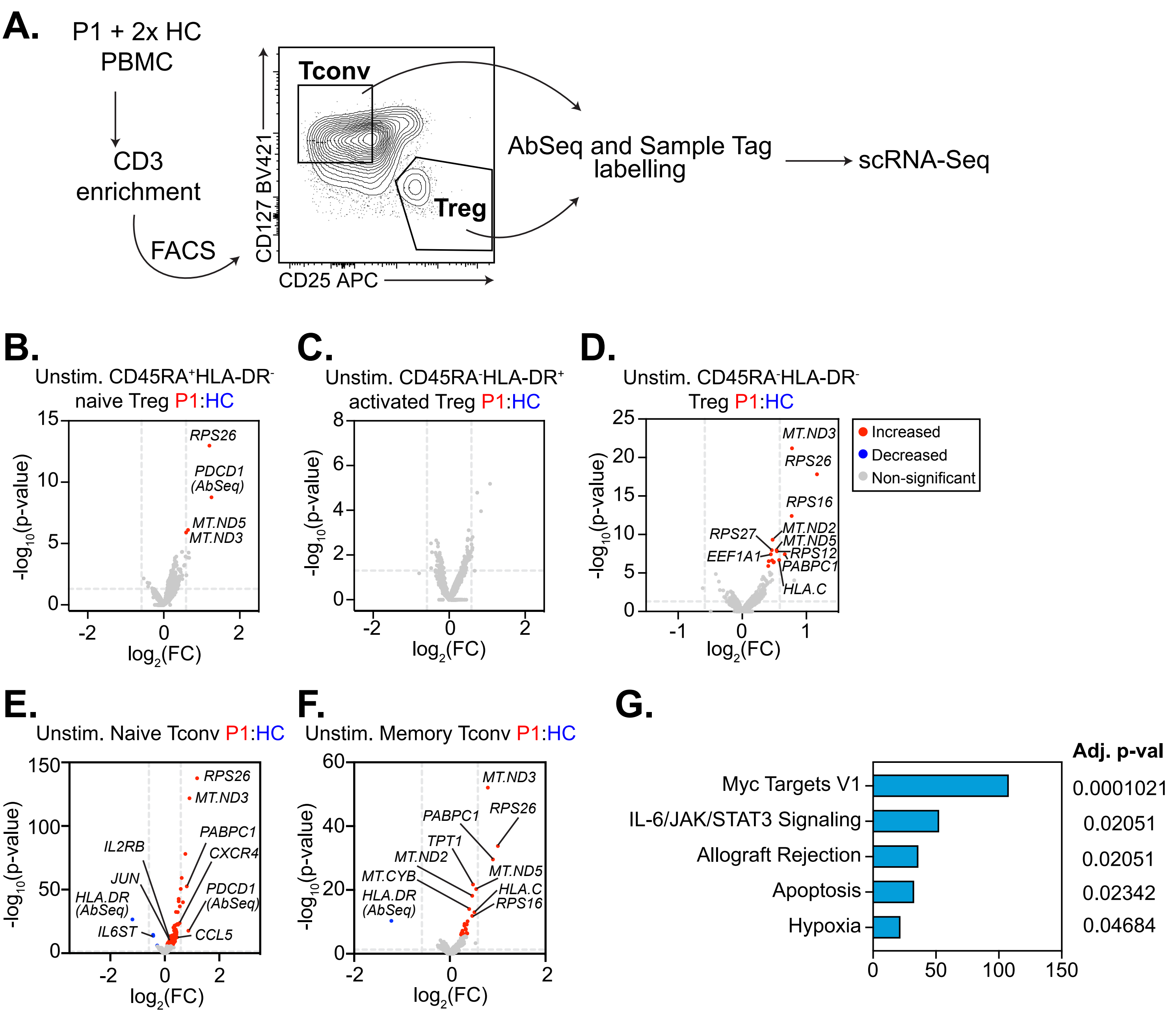

A.
P1 + 2x HC
PBMC
CD3
enrichment
FACS
Tconv
Treg
CD127 BV421
CD25 APC
AbSeq and Sample Tag labelling
scRNA-Seq
B.
Unstim. CD45RA+HLA-DR- naive Treg P1:HC
RPS26
PDCD1 (AbSeq)
MT.ND5
MT.ND3
C.
Unstim. CD45RA-HLA-DR+ activated Treg P1:HC
D.
Unstim. CD45RA-HLA-DR- Treg P1:HC
MT.ND3
RPS26
RPS16
RPS27
MT.ND2
MT.ND5
EEF1A1
RPS12
PABPC1
HLA.C
Increased
Decreased
Non-significant
E.
Unstim. Naive Tconv P1:HC
RPS26
MT.ND3
PABPC1
IL2RB
CXCR4
JUN
PDCD1 (AbSeq)
HLA.DR (AbSeq)
IL6ST
CCL5
F.
Unstim. Memory Tconv P1:HC
MT.ND3
PABPC1
RPS26
TPT1
MT.ND2
MT.CYB
MT.ND5
HLA.DR (AbSeq)
HLA.C
RPS16
-log10(p-value)
log2(FC)
G.
Adj. p-val
Myc Targets V1
IL-6/JAK/STAT3 Signaling
Allograft Rejection
Apoptosis
Hypoxia
0.0001021
0.02051
0.02051
0.02342
0.04684

### Figure S7

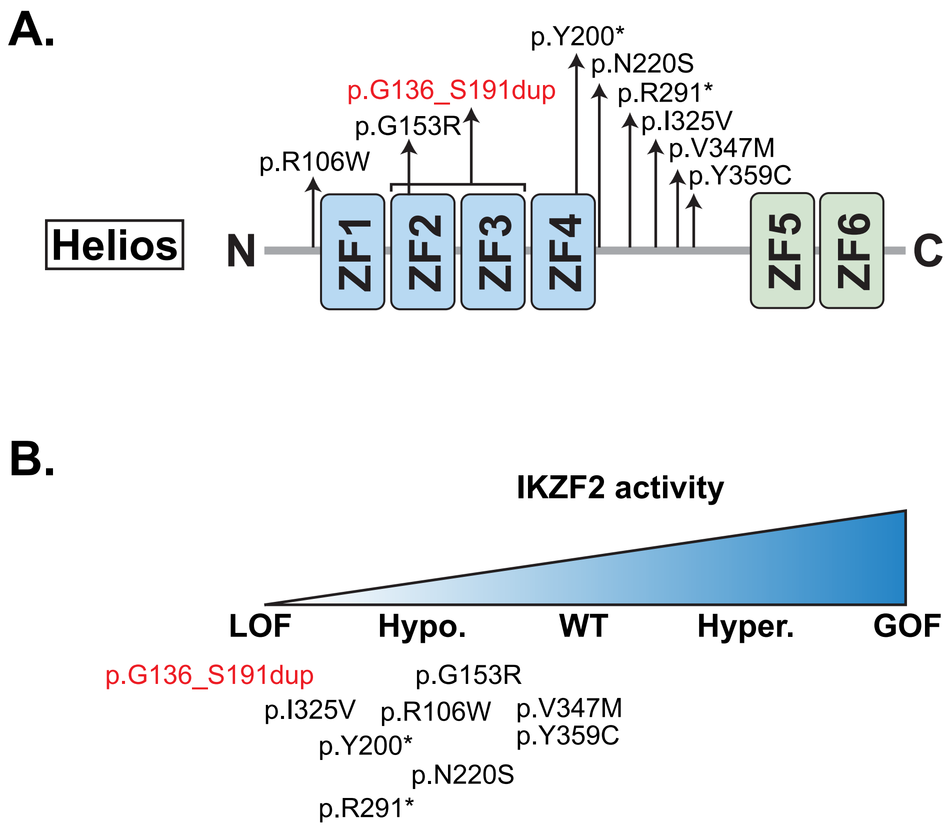
A.
p.Y200*
p.N220S
p.G136_S191dup
p.R291*
p.I325V
p.G153R
p.V347M
p.R106W
p.Y359C
Helios
N
ZF1
ZF2
ZF3
ZF4
ZF5
ZF6
C
B.
IKZF2 activity
LOF
Hypo.
WT
Hyper.
GOF
p.G136_S191dup
p.G153R
p.I325V
p.R106W
p.V347M
p.Y359C
p.Y200*
p.N220S
p.R291*

### Table S1

| **Unstimulated CD45RA$^{+}$HLA-DR$^{-}$ Tregs P1/HC** | | | | |
|---|---|---|---|---|
| Gene | Average $\log_2$(FC) | Patient % reads | Controls (n=2) % reads | Adjusted p-value |
| *RPS26* | 1.20410776 | 0.781 | 0.275 | 2.05E-09 |
| *CD279 (PD-1) AbSeq* | 1.25536754 | 0.938 | 0.941 | 3.32E-05 |
| *MT.ND5* | 0.63759536 | 1 | 0.985 | 0.01554611 |
| *MT.ND3* | 0.58915204 | 1 | 1 | 0.02481357 |
| **Stimulated CD45RA$^{+}$HLA-DR$^{-}$ Tregs P1/HC** | | | | |
| Gene | Average $\log_2$(FC) | Patient % reads | Controls (n=2) % reads | Adjusted p-value |
| *IL2* | 2.33734243 | 0.304 | 0.188 | 3.65E-08 |
| *CCR4* | 0.89037551 | 0.478 | 0.062 | 6.64E-05 |
| *CD279 (PD-1) AbSeq* | 1.52169856 | 1 | 0.987 | 0.00287593 |
| **Unstimulated CD45RA$^{-}$HLA-DR$^{+}$ Tregs P1/HC** | | | | |
| Gene | Average $\log_2$(FC) | Patient % reads | Controls (n=2) % reads | Adjusted p-value |
| — | — | — | — | — |
| **Stimulated CD45RA$^{-}$HLA-DR$^{+}$ Tregs P1/HC** | | | | |
| Gene | Average $\log_2$(FC) | Patient % reads | Controls (n=2) % reads | Adjusted p-value |
| *RPS3A* | 0.85306912 | 0.979 | 0.846 | 0.00072689 |
| *RPS4X* | 0.6578455 | 0.957 | 0.872 | 0.05296446 |
| **Unstimulated CD45RA$^{-}$HLA-DR$^{-}$ Tregs P1/HC** | | | | |
| Gene | Average $\log_2$(FC) | Patient % reads | Controls (n=2) % reads | Adjusted p-value |
| *MT.ND3* | 0.77616704 | 0.994 | 1 | 1.24E-17 |
| *RPS26* | 1.16556698 | 0.754 | 0.253 | 2.90E-14 |
| *RPS16* | 0.77245575 | 0.928 | 0.714 | 7.73E-09 |
| *MT.ND2* | 0.4733298 | 1 | 1 | 9.40E-06 |
| *MT.ND5* | 0.53517757 | 0.988 | 1 | 0.00021311 |
| *RPS27* | 0.46032408 | 0.994 | 0.978 | 0.00021528 |
| *RPS12* | 0.54074794 | 0.988 | 0.956 | 0.00028853 |
| *PABPC1* | 0.66441632 | 0.94 | 0.802 | 0.00073903 |
| *EEF1A1* | 0.44542665 | 1 | 0.978 | 0.00073941 |
| *HLA.C* | 0.57526163 | 0.832 | 0.725 | 0.00421473 |
| *RPS27A* | 0.46500459 | 1 | 0.945 | 0.00463577 |
| *TPT1* | 0.41422608 | 1 | 0.989 | 0.00576331 |
| *RPS23* | 0.4942834 | 0.976 | 0.89 | 0.0073653 |
| *RPS13* | 0.48931195 | 0.982 | 0.912 | 0.00807184 |
| *MT.CYB* | 0.40709176 | 0.994 | 0.989 | 0.02477644 |
| **Stimulated CD45RA$^{-}$HLA-DR$^{-}$ Tregs P1/HC** | | | | |
| Gene | Average $\log_2$(FC) | Patient % reads | Controls (n=2) % reads | Adjusted p-value |

| | | | | |
|---|---|---|---|---|
| *IL21* | -0.7570734 | 0.018 | 0.028 | 3.94E-12 |
| *RPS26* | 0.8831303 | 0.798 | 0.425 | 4.22E-11 |
| *MT.ND3* | 0.59101846 | 0.988 | 1 | 1.49E-06 |
| *RPS16* | 0.55000266 | 0.926 | 0.821 | 0.00265058 |
| *TPT1* | 0.34717656 | 1 | 1 | 0.0191753 |
| *HLA.C* | 0.51458739 | 0.859 | 0.708 | 0.02675433 |
| *RPS3A* | 0.43639648 | 1 | 0.991 | 0.03257976 |

### Table S2

| Unstimulated CD45RA$^+$ naïve Tconv P1/HC | | | | |
|---|---|---|---|---|
| Gene | Average $log_2$(FC) | Patient % reads | Controls (n=2) % reads | Adjusted p-value |
| *RPS26* | 1.18764709 | 0.835 | 0.251 | 4.85E-134 |
| *MT.ND3* | 0.9058343 | 1 | 0.998 | 3.14E-118 |
| *MT.ND5* | 0.75680139 | 0.995 | 0.994 | 1.54E-74 |
| *MT.ND2* | 0.61922654 | 0.997 | 1 | 1.16E-55 |
| *PABPC1* | 0.80699577 | 0.984 | 0.907 | 5.28E-49 |
| *MT.CYB* | 0.58768225 | 0.997 | 0.999 | 4.66E-47 |
| *MT.ND1* | 0.50909671 | 0.997 | 0.999 | 4.50E-39 |
| *MT.ATP6* | 0.51007248 | 1 | 0.999 | 3.25E-37 |
| *RPS16* | 0.66691202 | 0.938 | 0.78 | 1.53E-36 |
| *TPT1* | 0.54980831 | 1 | 0.996 | 3.06E-33 |
| *MT.CO1* | 0.40887775 | 1 | 1 | 6.28E-29 |
| *MT.CO3* | 0.47825594 | 1 | 1 | 1.10E-28 |
| *CD74 (HLA.DR) AbSeq* | -1.1867976 | 0.989 | 0.99 | 5.39E-23 |
| *CXCR4* | 0.54563294 | 0.622 | 0.325 | 3.82E-20 |
| *RPS20* | 0.46388552 | 0.992 | 0.946 | 1.99E-19 |
| *EEF1A1* | 0.48001202 | 1 | 0.989 | 4.41E-19 |
| *HLA.C* | 0.4851569 | 0.93 | 0.803 | 7.04E-19 |
| *RPS3A* | 0.41255559 | 0.995 | 0.974 | 1.11E-18 |
| *RPS29* | 0.35973859 | 1 | 0.998 | 3.79E-18 |
| *PDE4DIP* | 0.3503569 | 0.281 | 0.079 | 2.72E-17 |
| *MT.ND4* | 0.31957801 | 0.997 | 1 | 1.13E-16 |
| *RPL37* | 0.40383317 | 0.997 | 0.961 | 5.64E-15 |
| *RPS27A* | 0.34302648 | 0.997 | 0.992 | 3.18E-14 |
| *CD279 (PD-1) AbSeq* | 0.86647915 | 0.93 | 0.924 | 3.78E-14 |
| *RPS3* | 0.42223693 | 0.884 | 0.733 | 1.07E-13 |
| *RPL10A* | 0.42589663 | 0.911 | 0.833 | 4.75E-13 |
| *RPS7* | 0.3963419 | 0.938 | 0.809 | 5.78E-13 |
| *RPS21* | 0.40979573 | 0.938 | 0.841 | 6.79E-13 |
| *RPS4X* | 0.4262625 | 0.986 | 0.922 | 9.16E-13 |
| *RPL3* | 0.4017918 | 0.938 | 0.878 | 3.85E-12 |
| *RPL5* | 0.38228455 | 0.959 | 0.896 | 1.41E-11 |
| *RPL34* | 0.3530331 | 1 | 0.989 | 1.48E-11 |
| *NAP1L1* | 0.42743514 | 0.884 | 0.798 | 1.96E-11 |
| *MT.ATP8* | 0.35716449 | 0.962 | 0.919 | 6.70E-11 |
| *RPL23A* | 0.34477365 | 0.989 | 0.935 | 6.74E-11 |
| *IL6ST* | -0.4264239 | 0.203 | 0.432 | 7.26E-11 |
| *RPL37A* | 0.34709528 | 0.986 | 0.949 | 1.10E-10 |
| *MT.CO2* | 0.26563944 | 1 | 0.999 | 2.60E-10 |
| *LRRN3* | -0.4252349 | 0.119 | 0.322 | 3.82E-10 |
| *RPS23* | 0.35711025 | 0.984 | 0.948 | 4.55E-10 |

### Table S2

| | | | | |
|---|---|---|---|---|
| *MIAT* | 0.19162171 | 0.135 | 0.015 | 7.17E-10 |
| *RPL6* | 0.35780755 | 0.976 | 0.911 | 1.09E-09 |
| *RPS27* | 0.31529748 | 0.997 | 0.988 | 2.12E-09 |
| *RPS8* | 0.35502653 | 0.992 | 0.975 | 2.69E-09 |
| *CCL5* | 0.45402733 | 0.122 | 0.005 | 5.41E-09 |
| *MT.ND4L* | 0.31499827 | 0.978 | 0.949 | 3.29E-08 |
| *IL2RB* | 0.17705987 | 0.111 | 0.012 | 4.25E-08 |
| *RTKN2* | 0.25091521 | 0.146 | 0.023 | 9.96E-08 |
| *EEF1B2* | 0.37647498 | 0.846 | 0.722 | 2.00E-07 |
| *RPL27* | 0.31837102 | 0.962 | 0.894 | 3.46E-07 |
| *AHNAK* | 0.28709213 | 0.257 | 0.095 | 3.85E-07 |
| *DHRS7* | 0.20209788 | 0.168 | 0.055 | 4.94E-07 |
| *RPL23* | 0.31184961 | 0.986 | 0.941 | 8.63E-07 |
| *RPS12* | 0.30558841 | 0.978 | 0.964 | 9.44E-07 |
| *RPL10* | 0.31995662 | 0.924 | 0.848 | 9.88E-07 |
| *ZNF90* | 0.27707266 | 0.438 | 0.28 | 1.08E-06 |
| *RPL39* | 0.29374602 | 0.995 | 0.967 | 1.48E-06 |
| *NHSL2* | 0.28545998 | 0.53 | 0.367 | 1.76E-06 |
| *EVI2B* | 0.33094594 | 0.543 | 0.351 | 1.78E-06 |
| *CENPK* | 0.1636345 | 0.135 | 0.036 | 3.39E-06 |
| *RPL38* | 0.31004347 | 0.935 | 0.88 | 3.53E-06 |
| *RPS6* | 0.28380205 | 1 | 0.991 | 4.14E-06 |
| *VIM* | 0.36951853 | 0.681 | 0.536 | 7.22E-06 |
| *JUN* | 0.21211469 | 0.173 | 0.067 | 7.94E-06 |
| *SYNE2* | 0.37496256 | 0.446 | 0.268 | 8.67E-06 |
| *RPL32* | 0.28319164 | 0.992 | 0.973 | 9.29E-06 |
| *02-Sep* | 0.27278652 | 0.386 | 0.241 | 9.73E-06 |
| *CSTB* | 0.24111641 | 0.3 | 0.155 | 1.17E-05 |
| *ARL4C* | 0.33146474 | 0.551 | 0.417 | 1.26E-05 |
| *JAZF1* | 0.19199078 | 0.159 | 0.047 | 2.62E-05 |
| *RPS15A* | 0.30448236 | 0.943 | 0.87 | 3.33E-05 |
| *TUBB* | 0.29222949 | 0.427 | 0.263 | 3.97E-05 |
| *RPL27A* | 0.35245909 | 0.873 | 0.791 | 4.32E-05 |
| *RPL13A* | 0.27241921 | 0.989 | 0.958 | 4.75E-05 |
| *EEF2* | 0.29929198 | 0.532 | 0.349 | 5.12E-05 |
| *SESN3* | 0.29666692 | 0.265 | 0.147 | 0.0001036 |
| *RPL35A* | 0.25816695 | 0.968 | 0.954 | 0.00013276 |
| *CD84* | 0.11816618 | 0.084 | 0.015 | 0.00013383 |
| *LINC01871* | 0.1132009 | 0.07 | 0.006 | 0.00018274 |
| *PLP2* | 0.20814347 | 0.184 | 0.068 | 0.00018419 |
| *RPL30* | 0.25554187 | 0.995 | 0.967 | 0.0002914 |
| *CD44* | 0.28085557 | 0.719 | 0.597 | 0.00041173 |
| *CCNI* | 0.29227033 | 0.695 | 0.565 | 0.00053983 |
| *ITGA4* | 0.21645487 | 0.208 | 0.091 | 0.00071131 |
| *IL32* | 0.25704165 | 0.368 | 0.218 | 0.00083061 |

| | | | | |
|---|---|---|---|---|
| *TAB2* | 0.23547289 | 0.351 | 0.193 | 0.00094034 |
| *RPL19* | 0.27438138 | 0.822 | 0.73 | 0.00094311 |
| *RPS5* | 0.29907769 | 0.808 | 0.721 | 0.00097342 |
| *ACTG1* | 0.31031392 | 0.77 | 0.668 | 0.00098564 |
| *CR1* | 0.15772106 | 0.111 | 0.028 | 0.00101276 |
| *RPLP2* | 0.27362006 | 0.868 | 0.785 | 0.00108883 |
| *SLC25A53* | 0.09662145 | 0.065 | 0.005 | 0.00124382 |
| *PABPC3* | 0.16096878 | 0.197 | 0.091 | 0.00159045 |
| *CD2* | 0.28822297 | 0.549 | 0.445 | 0.00169603 |
| *RPLP1* | 0.2575776 | 0.903 | 0.827 | 0.00248375 |
| *RPL21* | 0.22764356 | 0.995 | 0.967 | 0.00250351 |
| *MACF1* | 0.25404093 | 0.295 | 0.183 | 0.0026387 |
| *RPL41* | 0.22209607 | 0.995 | 0.978 | 0.00265406 |
| *RPS13* | 0.22008659 | 0.997 | 0.982 | 0.00283929 |
| *RPL9* | 0.22827667 | 0.986 | 0.973 | 0.00297188 |
| *SPOCK2* | 0.27135439 | 0.651 | 0.538 | 0.00335293 |
| *RPL13* | 0.27413158 | 0.816 | 0.697 | 0.00363511 |
| *ANXA2* | 0.10179614 | 0.07 | 0.014 | 0.0038377 |
| *CLDND1* | 0.18054461 | 0.146 | 0.064 | 0.00531297 |
| *PFDN5* | 0.25561479 | 0.592 | 0.435 | 0.00547469 |
| *RPL12* | 0.29663665 | 0.868 | 0.735 | 0.00748031 |
| *RPL11* | 0.23705899 | 0.973 | 0.953 | 0.00934594 |
| *MYL12B* | 0.24102862 | 0.573 | 0.466 | 0.01137859 |
| *LEF1* | -0.2791661 | 0.503 | 0.608 | 0.01556475 |
| *RPL15* | 0.25246038 | 0.854 | 0.741 | 0.01680116 |
| *S100A4* | 0.14450502 | 0.111 | 0.041 | 0.01981441 |
| *RPL4* | 0.25523256 | 0.789 | 0.66 | 0.02561724 |
| *MYO9A* | 0.15146107 | 0.119 | 0.052 | 0.02658199 |
| *LDHA* | 0.20010635 | 0.332 | 0.214 | 0.02687985 |
| *SERPINB9* | 0.20540834 | 0.273 | 0.147 | 0.03192208 |
| *TBCD* | 0.08983582 | 0.086 | 0.025 | 0.03648794 |
| *ANXA1* | 0.22257962 | 0.259 | 0.147 | 0.0416569 |
| *RPL7* | 0.23760338 | 0.922 | 0.841 | 0.04625792 |
| *ZCCHC18* | 0.08348203 | 0.046 | 0.004 | 0.05331048 |
| *KLF6* | 0.21809127 | 0.281 | 0.177 | 0.0542021 |

**Stimulated CD45RA$^{+}$ naïve Tconv P1/HC**

| Gene | Average $log_2$(FC) | Patient % reads | Controls (n=2) % reads | Adjusted p-value |
|---|---|---|---|---|
| *RPS26* | 1.07510804 | 0.842 | 0.43 | 1.61E-72 |
| *MT.ND3* | 0.81265879 | 1 | 0.997 | 1.93E-57 |
| *CCL4* | 3.35267932 | 0.158 | 0.063 | 1.19E-46 |
| *RPS16* | 0.73852118 | 0.977 | 0.862 | 7.16E-31 |
| *CCL4L2* | 3.10429541 | 0.088 | 0.035 | 9.21E-29 |
| *B2M* | 0.47290926 | 1 | 0.999 | 2.63E-22 |
| *IL2* | 3.0415696 | 0.512 | 0.191 | 4.46E-22 |

### Table S2

| | | | | |
|---|---|---|---|---|
| *NDFIP2* | 0.62565985 | 0.591 | 0.272 | 5.81E-22 |
| *HSPA5* | 1.14864982 | 0.874 | 0.627 | 1.29E-21 |
| *LDHA* | 0.72694168 | 0.893 | 0.708 | 1.95E-21 |
| *RPL12* | 0.72767319 | 0.944 | 0.772 | 2.99E-20 |
| *HLA.C* | 0.61961593 | 0.893 | 0.74 | 3.44E-20 |
| *MT.ND5* | 0.51132604 | 0.991 | 0.976 | 5.79E-20 |
| *RPS23* | 0.51825583 | 1 | 0.966 | 2.09E-18 |
| *IL32* | 0.70567663 | 0.563 | 0.212 | 3.79E-18 |
| *IRF8* | 1.21888756 | 0.54 | 0.207 | 1.36E-17 |
| *TNFRSF9* | 0.9033837 | 0.4 | 0.07 | 1.54E-17 |
| *PTMA* | 0.4933372 | 1 | 0.995 | 1.79E-17 |
| *RAN* | 0.58851994 | 0.916 | 0.819 | 1.94E-17 |
| *HSP90AB1* | 0.53639726 | 1 | 0.987 | 1.99E-17 |
| *ODC1* | 0.63714795 | 0.693 | 0.407 | 7.59E-17 |
| *MT.CYB* | 0.44035863 | 0.995 | 0.997 | 1.59E-16 |
| *MT.ND2* | 0.40356237 | 1 | 1 | 2.12E-16 |
| *BCL2L11* | 0.60639601 | 0.363 | 0.108 | 2.66E-16 |
| *NPM1* | 0.53444938 | 0.995 | 0.984 | 2.97E-16 |
| *MIR155HG* | 0.77993904 | 0.981 | 0.904 | 1.30E-15 |
| *CD2* | 0.70941712 | 0.898 | 0.671 | 3.16E-15 |
| *FABP5* | 1.01295629 | 0.623 | 0.244 | 1.33E-14 |
| *CD48* | 0.65118101 | 0.693 | 0.457 | 3.49E-14 |
| *RPS6* | 0.46795979 | 1 | 0.993 | 3.83E-14 |
| *EGR1* | -0.9263666 | 0.549 | 0.732 | 4.32E-14 |
| *MT.CO1* | 0.44274492 | 1 | 1 | 4.53E-14 |
| *PGAM1* | 0.62518598 | 0.581 | 0.278 | 8.34E-14 |
| *PDIA6* | 0.57175931 | 0.516 | 0.219 | 9.53E-14 |
| *TPT1* | 0.42309298 | 1 | 1 | 1.07E-13 |
| *RPL10A* | 0.46714299 | 0.995 | 0.941 | 1.26E-13 |
| *TUBA1B* | 0.5406853 | 0.665 | 0.403 | 2.76E-13 |
| *RPL3* | 0.4888052 | 0.963 | 0.899 | 5.13E-13 |
| *RPL37* | 0.46649178 | 0.986 | 0.974 | 8.00E-13 |
| *HSPA8* | 0.6295723 | 0.823 | 0.627 | 1.12E-12 |
| *RPS29* | 0.39926115 | 1 | 0.999 | 1.30E-12 |
| *VIM* | 1.01838079 | 0.651 | 0.39 | 1.64E-12 |
| *YBX1* | 0.53055372 | 0.949 | 0.854 | 2.33E-12 |
| *RPL6* | 0.40043821 | 1 | 0.988 | 2.78E-12 |
| *APOBEC3G* | 0.26512334 | 0.121 | 0.01 | 3.63E-12 |
| *PPIA* | 0.45755251 | 0.977 | 0.949 | 5.18E-12 |
| *RPL41* | 0.40876842 | 1 | 0.984 | 9.35E-12 |
| *EEF1A1* | 0.37607345 | 1 | 0.998 | 1.10E-11 |
| *IFNG* | 2.42783618 | 0.205 | 0.069 | 1.79E-11 |
| *CISH* | 0.27213271 | 0.191 | 0.039 | 2.07E-11 |
| *NCL* | 0.50475565 | 0.94 | 0.892 | 2.17E-11 |
| *IER3* | 0.82091044 | 0.642 | 0.371 | 2.71E-11 |

### Table S2

| | | | | |
|---|---|---|---|---|
| *CYTOR* | 0.56174918 | 0.344 | 0.091 | 2.90E-11 |
| *TBX21* | 0.54564065 | 0.4 | 0.112 | 3.91E-11 |
| *RPS12* | 0.44211573 | 1 | 0.975 | 4.21E-11 |
| *MT.ND1* | 0.37084613 | 1 | 0.998 | 4.40E-11 |
| *RPS3A* | 0.42164553 | 1 | 0.979 | 4.80E-11 |
| *SLC7A5* | 0.56118287 | 0.647 | 0.442 | 6.48E-11 |
| *CCL3* | 1.08068476 | 0.065 | 0.009 | 7.27E-11 |
| *RPS27A* | 0.39466837 | 1 | 0.991 | 7.79E-11 |
| *RPS27* | 0.39432235 | 1 | 0.992 | 9.05E-11 |
| *APOBEC3C* | 0.31379398 | 0.121 | 0.011 | 9.52E-11 |
| *ATXN1* | 0.4125093 | 0.242 | 0.066 | 1.27E-10 |
| *RPL7* | 0.443437 | 0.981 | 0.917 | 1.37E-10 |
| *TRMT112* | 0.41456017 | 0.526 | 0.281 | 2.31E-10 |
| *RPLP1* | 0.43260384 | 0.958 | 0.905 | 2.99E-10 |
| *PRNP* | 0.56212877 | 0.665 | 0.458 | 4.05E-10 |
| *RPS3* | 0.45277348 | 0.926 | 0.819 | 4.68E-10 |
| *RPL37A* | 0.40404607 | 1 | 0.97 | 7.28E-10 |
| *RPS4X* | 0.48130723 | 0.995 | 0.92 | 7.48E-10 |
| *SAMSN1* | 0.56149574 | 0.544 | 0.313 | 9.96E-10 |
| *ZNF90* | 0.40733656 | 0.577 | 0.325 | 1.81E-09 |
| *NAMPT* | 0.77328115 | 0.735 | 0.52 | 2.38E-09 |
| *CFLAR* | 0.6137928 | 0.823 | 0.591 | 3.31E-09 |
| *ATP1B3* | 0.50893291 | 0.581 | 0.35 | 4.53E-09 |
| *NAP1L1* | 0.40336333 | 0.991 | 0.949 | 5.23E-09 |
| *KCNK5* | 0.16717097 | 0.116 | 0.015 | 7.36E-09 |
| *ENO1* | 0.45880546 | 0.935 | 0.848 | 7.50E-09 |
| *CYCS* | 0.4680815 | 0.847 | 0.77 | 8.93E-09 |
| *MT.ATP6* | 0.34177897 | 1 | 0.999 | 9.94E-09 |
| *STK24* | 0.31489917 | 0.247 | 0.083 | 1.53E-08 |
| *RPS21* | 0.41361322 | 0.977 | 0.896 | 1.54E-08 |
| *CD82* | 0.38683963 | 0.293 | 0.109 | 1.62E-08 |
| *GAPDH* | 0.46044322 | 0.553 | 0.308 | 1.75E-08 |
| *RPS28* | 0.43413093 | 0.916 | 0.806 | 2.05E-08 |
| *TNF* | 1.08581562 | 0.409 | 0.174 | 2.58E-08 |
| *RPS24* | 0.38580888 | 0.981 | 0.964 | 2.94E-08 |
| *PKM* | 0.48359244 | 0.907 | 0.826 | 6.83E-08 |
| *ZFP36L1* | 0.69014472 | 0.805 | 0.731 | 8.04E-08 |
| *MDFIC* | 0.53323077 | 0.6 | 0.355 | 1.13E-07 |
| *IRF2BP2* | -0.5322223 | 0.349 | 0.582 | 1.13E-07 |
| *LAPTM4B* | 0.21451445 | 0.172 | 0.036 | 1.38E-07 |
| *LRIG1* | 0.22438831 | 0.158 | 0.034 | 1.54E-07 |
| *TMSB4X* | -0.603295 | 0.856 | 0.93 | 1.68E-07 |
| *RPS20* | 0.35562436 | 0.995 | 0.98 | 2.05E-07 |
| *RGCC* | 0.60303062 | 0.609 | 0.425 | 2.28E-07 |
| *RPL5* | 0.36230651 | 0.991 | 0.968 | 2.57E-07 |

### Table S2

| | | | | |
|---|---|---|---|---|
| *SACS* | 0.48274329 | 0.577 | 0.309 | 3.88E-07 |
| *CDK6* | 0.59979788 | 0.563 | 0.337 | 4.65E-07 |
| *ARHGDIB* | -0.4951369 | 0.172 | 0.411 | 5.20E-07 |
| *PARK7* | 0.37081218 | 0.507 | 0.29 | 5.92E-07 |
| *MTRNR2L8* | 0.33501939 | 0.521 | 0.365 | 6.40E-07 |
| *RPL34* | 0.3513544 | 1 | 0.986 | 1.34E-06 |
| *NBEAL1* | 0.40682923 | 0.656 | 0.526 | 1.44E-06 |
| *RPL11* | 0.37692217 | 0.995 | 0.946 | 1.54E-06 |
| *PPP1CC* | 0.37553109 | 0.447 | 0.283 | 2.82E-06 |
| *CD3D* | 0.41652727 | 0.581 | 0.408 | 2.97E-06 |
| *XBP1* | 0.43668003 | 0.623 | 0.342 | 3.12E-06 |
| *RPS8* | 0.38267574 | 1 | 0.968 | 3.53E-06 |
| *RPL31* | 0.35343918 | 1 | 0.977 | 3.99E-06 |
| *LRRFIP1* | 0.41110562 | 0.54 | 0.346 | 4.42E-06 |
| *TUBA1C* | 0.31104468 | 0.367 | 0.18 | 4.57E-06 |
| *TUBB* | 0.42508708 | 0.488 | 0.274 | 4.68E-06 |
| *CD3E* | 0.41796416 | 0.563 | 0.384 | 6.26E-06 |
| *NR4A1* | -0.677923 | 0.414 | 0.555 | 7.74E-06 |
| *SEMA4D* | 0.27467509 | 0.247 | 0.099 | 8.25E-06 |
| *RPS13* | 0.30882593 | 1 | 0.988 | 8.69E-06 |
| *COPRS* | 0.19146662 | 0.116 | 0.028 | 9.78E-06 |
| *CD84* | 0.34669049 | 0.177 | 0.047 | 1.05E-05 |
| *RPL27* | 0.34874057 | 0.972 | 0.922 | 1.41E-05 |
| *RPL39* | 0.32680061 | 1 | 0.982 | 1.43E-05 |
| *CALR* | 0.43681218 | 0.53 | 0.351 | 1.49E-05 |
| *GEM* | -0.740959 | 0.363 | 0.488 | 1.55E-05 |
| *F5* | 0.29543444 | 0.135 | 0.031 | 1.77E-05 |
| *CCT8* | 0.37178869 | 0.609 | 0.425 | 2.60E-05 |
| *EEF1B2* | 0.35303849 | 0.972 | 0.938 | 2.85E-05 |
| *RPS7* | 0.35166741 | 0.981 | 0.912 | 3.43E-05 |
| *TOMM7* | 0.39634813 | 0.753 | 0.579 | 3.97E-05 |
| *HNRNPAB* | 0.38621372 | 0.693 | 0.565 | 4.04E-05 |
| *SNRPF* | 0.31636246 | 0.447 | 0.247 | 4.69E-05 |
| *BTF3* | 0.34356033 | 0.926 | 0.761 | 4.99E-05 |
| *RPL21* | 0.30437834 | 0.995 | 0.991 | 5.40E-05 |
| *EXOC2* | 0.29481365 | 0.335 | 0.139 | 5.72E-05 |
| *BX890604.2* | -0.3388955 | 0.051 | 0.254 | 6.02E-05 |
| *RPL23A* | 0.30918301 | 0.986 | 0.955 | 6.33E-05 |
| *CD200* | 0.89590263 | 0.6 | 0.261 | 6.67E-05 |
| *SLAMF1* | 0.55837826 | 0.488 | 0.204 | 6.88E-05 |
| *NME1* | 0.38015908 | 0.586 | 0.397 | 0.00010134 |
| *INSIG1* | 0.74065541 | 0.512 | 0.289 | 0.00010543 |
| *SGPP2* | 0.16349503 | 0.084 | 0.011 | 0.00011276 |
| *CRTAM* | 0.67438848 | 0.149 | 0.011 | 0.00011366 |
| *RPL38* | 0.34675142 | 0.977 | 0.896 | 0.00012027 |

### Table S2

| | | | | |
|---|---|---|---|---|
| *RBPJ* | 0.44136317 | 0.414 | 0.268 | 0.00012101 |
| *CHST11* | 0.18164862 | 0.144 | 0.03 | 0.00012232 |
| *RPL14* | 0.34543223 | 0.935 | 0.898 | 0.00012973 |
| *CCL3L1* | 0.42496415 | 0.051 | 0.003 | 0.0001332 |
| *HSP90B1* | 0.47328083 | 0.674 | 0.575 | 0.00015346 |
| *RPL32* | 0.31413776 | 1 | 0.98 | 0.00016342 |
| *SMCHD1* | -0.4256057 | 0.27 | 0.459 | 0.00018858 |
| *ATM* | -0.3608168 | 0.135 | 0.332 | 0.00028412 |
| *CERS2* | 0.26429921 | 0.279 | 0.124 | 0.0003234 |
| *RPL4* | 0.32460523 | 0.949 | 0.872 | 0.00036905 |
| *RPS15A* | 0.33150826 | 0.949 | 0.903 | 0.00039263 |
| *VMP1* | 0.4125161 | 0.553 | 0.346 | 0.00041234 |
| *SPRY1* | 0.61283754 | 0.349 | 0.085 | 0.0004614 |
| *RPLP2* | 0.34951443 | 0.888 | 0.8 | 0.00046634 |
| *SPCS2* | 0.28908392 | 0.349 | 0.219 | 0.0004733 |
| *GLUL* | 0.17866836 | 0.14 | 0.038 | 0.00048696 |
| *DNAJC3* | 0.28708093 | 0.209 | 0.104 | 0.00058476 |
| *PFKP* | 0.2506796 | 0.233 | 0.095 | 0.00060628 |
| *CLEC2B* | -0.3616149 | 0.107 | 0.264 | 0.00070804 |
| *DDX21* | 0.32267282 | 0.986 | 0.947 | 0.00078135 |
| *TFRC* | 0.38707592 | 0.651 | 0.461 | 0.00080741 |
| *CREM* | -0.4991119 | 0.633 | 0.726 | 0.0008145 |
| *PUS7* | 0.31829051 | 0.34 | 0.172 | 0.00086154 |
| *TAGAP* | 0.44694174 | 0.484 | 0.284 | 0.0008962 |
| *TOP1* | 0.36153672 | 0.651 | 0.514 | 0.00092775 |
| *MT.CO3* | 0.28093207 | 1 | 0.997 | 0.00107869 |
| *RPS11* | 0.34289459 | 0.809 | 0.701 | 0.0011362 |
| *MIR4435.2HG* | 0.21423761 | 0.13 | 0.023 | 0.00139046 |
| *ABLIM1* | -0.3560797 | 0.13 | 0.304 | 0.00147939 |
| *MTRNR2L12* | 0.32687056 | 0.902 | 0.816 | 0.00170657 |
| *GBP5* | 0.71998895 | 0.66 | 0.385 | 0.00230477 |
| *ADAM19* | 0.19885388 | 0.13 | 0.032 | 0.00238631 |
| *RRP1B* | 0.24589454 | 0.307 | 0.145 | 0.00248421 |
| *NIPAL4* | 0.08830141 | 0.06 | 0.006 | 0.00256035 |
| *CD226* | 0.2197003 | 0.107 | 0.022 | 0.00258651 |
| *BTG2* | -0.4211603 | 0.753 | 0.826 | 0.00267294 |
| *RPS5* | 0.36178066 | 0.907 | 0.808 | 0.00281864 |
| *NOP56* | 0.34591898 | 0.623 | 0.451 | 0.00283572 |
| *RPL19* | 0.31757026 | 0.884 | 0.787 | 0.00289839 |
| *SLC25A5* | 0.30735295 | 0.66 | 0.559 | 0.00295012 |
| *PPP1R18* | 0.3315262 | 0.423 | 0.267 | 0.00297613 |
| *AMIGO2* | 0.28320859 | 0.247 | 0.125 | 0.00325212 |
| *FRMD4B* | 0.1170881 | 0.065 | 0.006 | 0.00350441 |
| *CCT2* | 0.33742868 | 0.823 | 0.677 | 0.00511971 |
| *EGR3* | -0.5618186 | 0.177 | 0.296 | 0.00547038 |

### Table S2

| | | | | |
|---|---|---|---|---|
| *RPL27A* | 0.38036642 | 0.902 | 0.79 | 0.00558135 |
| *RPL23* | 0.27876981 | 0.995 | 0.974 | 0.00562688 |
| *ZEB2* | 0.0968041 | 0.051 | 0.005 | 0.00633743 |
| *SEC61G* | 0.26927243 | 0.391 | 0.196 | 0.00639788 |
| *NCOA7* | 0.40717116 | 0.502 | 0.345 | 0.00681199 |
| *XCL2* | 0.33588999 | 0.033 | 0.002 | 0.00682642 |
| *HSPH1* | 0.35911584 | 0.628 | 0.464 | 0.00696678 |
| *TNFAIP8* | 0.4458074 | 0.833 | 0.579 | 0.00727415 |
| *NCS1* | 0.12585314 | 0.093 | 0.015 | 0.00736033 |
| *STAT4* | 0.28588064 | 0.26 | 0.124 | 0.00773559 |
| *GBP2* | 0.42106527 | 0.87 | 0.733 | 0.00897673 |
| *MSMO1* | 0.2848661 | 0.27 | 0.156 | 0.00919553 |
| *MALAT1* | -0.3236432 | 0.981 | 0.988 | 0.00969074 |
| *CALM2* | 0.30691484 | 0.66 | 0.523 | 0.00986095 |
| *PDCD4* | -0.3236357 | 0.195 | 0.366 | 0.01007615 |
| *NOLC1* | 0.33893961 | 0.833 | 0.744 | 0.01049243 |
| *STAMBPL1* | 0.12261457 | 0.07 | 0.01 | 0.0153637 |
| *SFXN1* | 0.50655846 | 0.558 | 0.314 | 0.01726372 |
| *CD44* | 0.33445664 | 0.981 | 0.936 | 0.01826508 |
| *SEMA7A* | 0.18211995 | 0.186 | 0.063 | 0.02059144 |
| *RPL9* | 0.24723599 | 0.995 | 0.989 | 0.0227567 |
| *HSPD1* | 0.36172782 | 0.809 | 0.618 | 0.02396607 |
| *RNF19A* | 0.39187594 | 0.409 | 0.231 | 0.02429099 |
| *CHSY1* | 0.38559816 | 0.595 | 0.415 | 0.02433046 |
| *PLEK* | 0.17436544 | 0.037 | 0.002 | 0.0244659 |
| *EIF4E* | 0.2592671 | 0.391 | 0.234 | 0.02582428 |
| *CSF2* | 0.15306881 | 0.042 | 0.003 | 0.02594697 |
| *GPR171* | 0.58065997 | 0.456 | 0.257 | 0.02677023 |
| *RPL36AL* | 0.27173131 | 0.595 | 0.439 | 0.02752894 |
| *WHAMM* | -0.2695546 | 0.074 | 0.208 | 0.02777574 |
| *RPLP0* | 0.28823561 | 0.623 | 0.49 | 0.02924261 |
| *NHSL2* | 0.25687144 | 0.665 | 0.539 | 0.02982679 |
| *PSMB2* | 0.25151863 | 0.386 | 0.27 | 0.03019139 |
| *LTA* | 0.26824217 | 0.149 | 0.06 | 0.03104481 |
| *KPNB1* | 0.36181443 | 0.73 | 0.604 | 0.03168615 |
| *G3BP1* | 0.32423621 | 0.791 | 0.67 | 0.03188377 |
| *POLR1B* | 0.19941859 | 0.237 | 0.124 | 0.03243354 |
| *CFL1* | 0.31882112 | 0.842 | 0.74 | 0.03271519 |
| *SERBP1* | 0.29556895 | 0.795 | 0.728 | 0.03274823 |
| *EPOP* | 0.22397143 | 0.237 | 0.122 | 0.03279995 |
| *TPI1* | 0.27626154 | 0.316 | 0.197 | 0.03304489 |
| *RRAS2* | 0.16501501 | 0.126 | 0.052 | 0.03576515 |
| *SP140* | 0.17386401 | 0.144 | 0.044 | 0.03674321 |
| *PHLDA1* | 0.53364129 | 0.614 | 0.42 | 0.03688378 |
| *PSME3* | 0.2968398 | 0.642 | 0.476 | 0.03738256 |

| | | | | |
|---|---|---|---|---|
| *PAICS* | 0.30542563 | 0.684 | 0.53 | 0.03837203 |
| *SLC16A1* | 0.2742894 | 0.381 | 0.237 | 0.04091637 |
| *BCL2L1* | 0.17261846 | 0.158 | 0.057 | 0.04190908 |
| *PA2G4* | 0.31205763 | 0.712 | 0.565 | 0.04232952 |
| *GRPEL1* | 0.30972028 | 0.493 | 0.341 | 0.04300741 |
| *SLAMF7* | 0.19498773 | 0.06 | 0.008 | 0.04361495 |
| *RPL13A* | 0.27270066 | 0.986 | 0.952 | 0.04744998 |
| *VDR* | 0.06523402 | 0.051 | 0.005 | 0.04924696 |
| **Unstimulated CD45RA$^-$ memory Tconv P1/HC** | | | | |
| Gene | Average log$_2$(FC) | Patient % reads | Controls (n=2) % reads | Adjusted p-value |
| *MT.ND3* | 0.79664877 | 0.996 | 0.995 | 1.68E-48 |
| *RPS26* | 1.00455673 | 0.731 | 0.281 | 3.42E-30 |
| *PABPC1* | 0.90110174 | 0.967 | 0.932 | 5.39E-26 |
| *TPT1* | 0.48490223 | 1 | 1 | 3.83E-18 |
| *MT.ND5* | 0.55081941 | 0.986 | 0.977 | 1.05E-16 |
| *MT.ND2* | 0.46906905 | 1 | 0.995 | 1.35E-14 |
| *MT.CYB* | 0.40593888 | 0.998 | 1 | 1.88E-10 |
| *HLA.C* | 0.5152003 | 0.934 | 0.76 | 1.88E-09 |
| *RPS16* | 0.46819457 | 0.938 | 0.842 | 2.58E-08 |
| *CD74 (HLA.DR) AbSeq* | -1.2202097 | 0.971 | 0.982 | 9.84E-07 |
| *RPS3A* | 0.37172433 | 0.992 | 0.973 | 1.30E-06 |
| *MT.ATP6* | 0.3071909 | 0.998 | 0.995 | 8.07E-06 |
| *RPS12* | 0.35611542 | 0.99 | 0.968 | 1.35E-05 |
| *MT.ND1* | 0.30639035 | 0.998 | 0.991 | 2.81E-05 |
| *EEF1A1* | 0.33065971 | 0.998 | 0.995 | 0.00028042 |
| *RPL37* | 0.3345126 | 0.994 | 0.982 | 0.00054379 |
| *MT.CO2* | 0.26659362 | 0.992 | 0.995 | 0.00118392 |
| *MT.ND4* | 0.26190779 | 0.998 | 1 | 0.00260473 |
| *RPS27A* | 0.26619792 | 1 | 0.995 | 0.00316743 |
| *RPL10A* | 0.35026828 | 0.896 | 0.837 | 0.00599892 |
| *RPS29* | 0.24547446 | 1 | 0.995 | 0.00657313 |
| *S1PR1* | 0.36404771 | 0.6 | 0.489 | 0.00863636 |
| *RPS23* | 0.2919371 | 0.99 | 0.964 | 0.01526154 |
| *MT.CO1* | 0.23379154 | 1 | 1 | 0.01880477 |
| *MT.ND4L* | 0.30343803 | 0.952 | 0.937 | 0.04017553 |
| **Stimulated CD45RA$^-$ memory Tconv P1/HC** | | | | |
| Gene | Average log$_2$(FC) | Patient % reads | Controls (n=2) % reads | Adjusted p-value |
| *RPS26* | 0.87629372 | 0.83 | 0.486 | 1.09E-27 |
| *MT.ND3* | 0.63313952 | 1 | 0.996 | 3.50E-26 |
| *HLA.C* | 0.70253517 | 0.935 | 0.78 | 1.98E-18 |
| *TUBB* | 0.63535096 | 0.514 | 0.255 | 6.29E-10 |
| *RPS16* | 0.46448734 | 0.971 | 0.934 | 2.83E-07 |

#### Table S2

| | | | | |
|---|---|---|---|---|
| *TPT1* | 0.36089467 | 1 | 1 | 1.36E-06 |
| *NDFIP2* | 0.47334677 | 0.493 | 0.297 | 2.32E-05 |
| *MT.ND2* | 0.32308847 | 1 | 1 | 3.88E-05 |
| *IL32* | 0.53343542 | 0.446 | 0.239 | 4.05E-05 |
| *RPS27A* | 0.32283712 | 0.996 | 0.985 | 7.23E-05 |
| *STARD4* | 0.51389348 | 0.475 | 0.263 | 0.00012494 |
| *MT.CYB* | 0.31803214 | 1 | 0.996 | 0.00044778 |
| *HSPA5* | 0.64199378 | 0.837 | 0.73 | 0.00052562 |
| *TUBA1B* | 0.44560096 | 0.685 | 0.444 | 0.00098888 |
| *B2M* | 0.26924664 | 1 | 1 | 0.0025367 |
| *IL3* | 0.26863353 | 0.007 | 0.012 | 0.00390776 |
| *RPS12* | 0.29494959 | 1 | 1 | 0.01418338 |
| *PPIA* | 0.30882176 | 0.989 | 0.961 | 0.01846865 |
| *CYSLTR1* | 0.37344481 | 0.254 | 0.097 | 0.0230222 |
| *RPS11* | 0.34909443 | 0.826 | 0.722 | 0.02322863 |
| *RPS27* | 0.24988588 | 1 | 0.996 | 0.04939569 |

### Table S3

| Helios variant | p.Arg106Trp | p.Gly136_Ser191dup | p.Gly153Arg | p.Tyr200* (Daughter) | p.Tyr200* (Father) | p.Asn220Ser | p.Arg291* | p.Ile325Val | p.Val347Met | p.Tyr359Cys (Daughter) | p.Tyr359Cys (Mother) |
|---|---|---|---|---|---|---|---|---|---|---|---|
| Zygosity | Het. | Het. | Het. | Het. | Het. | Het. | Het. | Homo. | Het. | Het. | Het. |
| Consanguinity | — | — | — | ND | ND | ND | — | + | — | — | — |
| Functional impact | LOF* | DN | DN | LOF* | LOF* | LOF* | LOF* | LOF | None | None | None |
| Impact on protein expression | None | ↓ | None | Truncated | Truncated | None | Truncated | None | None | None | None |
| Developmental abnormalities | — | + | + | — | — | — | — | + | — | — | — |
| Clinical phenotype | | | | | | | | | | | |
| Infections | + | + | + | + | + | — | — | + | + | — | — |
| Mononucleosis | + (EBV) | — | — | — | — | — | — | — | — | — | — |
| Recurrent (Sino)pulmonary/otitis media | + | + | + | + | + | — | — | + | + | — | — |
| Recurrent *C. albicans* (oral/vaginal) | — | — | — | + | + | — | — | — | — | — | — |
| Recurrent diarrhea | — | — | — | + | — | — | — | — | + | — | — |
| Recurrent stomatitis | — | — | — | — | — | — | — | — | + | — | — |
| Serositis | | | | | | | | | + | — | — |
| Lymphoma | — | — | — | — | + | — | — | — | — | — | — |
| (Focal segmental) glomerulonephritis | — | — | — | — | — | — | — | — | + | — | + |
| Immune phenotype | | | | | | | | | | | |
| CD3 | ↑ | ↓ | ↓ | Normal | ↓ | Normal | Normal | ↑ | ↓ | ND | ND |
| CD4 | ↑ | ↑ | ↓ | Normal | Normal | Normal | ↓ | ↓ | ↓ | ↓ | ↓ |
| Naive | Normal | ↓ | ND | Normal | ↓ | ND | Normal | Normal | ↓ | Normal | Normal |
| EM | Normal | Normal | ND | Normal | ↑ | ND | Normal | Normal | ↑ | ↓ | Normal |
| CM | Normal | ↑ | ND | ↑ | ↑ | ND | Normal | Normal | Normal | Normal | Normal |
| TEMRA | Normal | Normal | ND | Normal | Normal | ND | ↓ | Normal | Normal | Normal | Normal |
| Tregs | Normal | Normal | ND | ↓ | ↓ | ND | Normal | ↓ | ↑ | Normal | Normal |
| $T_H1$ | ND | ↑ | ND | ↑ | ↑ | ND | ND | Normal | ND | ND | ND |
| $T_H2$ | ND | ↑ | ND | ND | ND | ND | ND | Normal | ND | ND | ND |
| $T_H9$ | ND | ↑ | ND | ND | ND | ND | ND | ND | ND | ND | ND |
| $T_H17$ | ND | ↑ | ND | ND | ND | ND | ND | Normal | ND | ND | ND |
| $cT_{FH}$ | ND | ↑ | ND | ↓ | ↓ | ND | ND | Normal | ND | ND | ND |
| $T_H22$ | ND | Normal | ND | ND | ND | ND | ND | ND | ND | ND | ND |
| CD8 | ↑ | ↓ | ↓ | ↑ | ↑ | ↓ | Normal | ↑ | Normal | ↑ | ↑ |
| Naive | Normal | ↓ | ND | ↓ | ↓ | Normal | Normal | ↓ | ↓ | Normal | Normal |
| EM | Normal | Normal | ND | Normal | Normal | Normal | Normal | ↑ | ↑ | Normal | Normal |
| CM | Normal | ↑ | ND | ↑ | Normal | Normal | Normal | Normal | Normal | Normal | Normal |
| TEMRA | Normal | Normal | ND | ↑ | ↑ | Normal | Normal | Normal | ↓ | ↓ | ↓ |
| $CD4^-CD8^-\gamma\delta^+$ | ND | ND | ND | ↓ | ↓ | ND | ND | ↑ | ND | ND | ND |
| $CD4^-CD8^-\alpha\beta^+$ | ND | ND | ND | ↓ | ↓ | ND | ND | ↓ | ND | ND | ND |
| MAIT | ND | ND | ND | ↓ | ↓ | ND | ND | ↓ | ND | ND | ND |
| NK | Normal | ↑ | ND | Normal | ↑ | ↓ | ↓ | ↓ | Normal | Normal | ↓ |
| CD19 | ↑ | Normal | ↓ | Normal | Normal | Normal | Normal | ↓ | ND | ND | ND |
| Naive | Normal | Normal | ND | Normal | ↑ | Normal | Normal | ↑ | ND | ND | ND |
| Non-switched memory | ↑ | ↑ | ND | Normal | ↓ | ↑ | Normal | ↓ | ND | ND | ND |
| Switched memory | Normal | Normal | ND | ↓ | Normal | Normal | Normal | ↓ | ND | ↓ | Normal |
| Transitional | ND | ↓ | ND | ↑ | ↑ | ND | ND | ↑ | ND | ND | ND |
| Plasmablast | ND | Normal | ND | ↓ | ↓ | ND | ND | ↓ | ND | ND | ND |
| Autoantibodies | — | + | ND | + | + | — | + | — | — | + | + |
| Immunoglobulins | | | | | | | | | | | |

### Table S3

| | | | | | | | | | | | |
|---|---|---|---|---|---|---|---|---|---|---|---|
| IgG | Normal | IVIG | ↓ | ↓ | Normal | ND | ↑ | ↓ | ↓ | ND | ND |
| IgA | ↓ | Normal | Normal | Normal | Normal | ND | ND | ↓ | ↓ | ND | ND |
| IgM | Normal | Normal | Normal | ↓ | Normal | ND | ND | ↓ | ↓ | ND | ND |
| IgE | ND | ↑ | ND | ↑ | Normal | ND | ND | Normal | ND | ND | ND |
| Vaccine responses | ND | Normal | ↓ | ↓ | Normal | ND | ND | ↓ | ND | ND | ND |
| Diptheria | ND | Normal | ND | ↓ | Normal | ND | ND | ↓ | ND | ND | ND |
| Tetanus | ND | Normal | ↓ | ↓ | Normal | ND | ND | ↓ | ND | ND | ND |
| *S. pneumoniae* | ND | ND | ↓ | ↓ | Normal | ND | ND | ND | ND | ND | ND |
| *H. influenzae* | ND | ND | ↓ | ND | Normal | ND | ND | ND | ND | ND | ND |
| Immune dysregulation/ autoimmunity | + | + | — | + | + | + | + | + | + | + | + |
| HLH | + | — | — | — | — | — | — | — | + | — | — |
| Still’s disease/arthritis/joint pain | + | — | — | — | — | — | + | + | + | — | — |
| AIHA | — | + | — | — | — | + (Evan’s) | — | — | — | — | — |
| ITP | — | — | — | — | — | + (Evan’s) | + | — | — | + | — |
| Eczema | — | + | — | — | — | — | — | — | — | — | — |
| Lichen planus | — | — | — | — | + | — | — | — | — | — | — |
| Chronic lymphadenopathy | — | — | — | + | + | — | — | — | — | — | — |
| Vitiligo | — | — | — | + | — | — | — | — | — | — | — |
| Colitis | — | — | — | — | + | — | — | — | — | — | — |
| Hypothyroidism | — | — | — | + | + | — | — | + | — | — | — |
| SLE | — | — | — | — | — | — | + | — | — | + | + |
| Reference | Shahin | Lu | Mohajeri | Hetemaki | Hetemaki | Shahin | Shahin | Shahin | Shahin | Shahin | Shahin |

### Table S4

| Phenotype | Frequency |
|---|---|
| Heterozygous *IKZF2* variant | 10/13 (77%) |
| Recurrent sinopulmonary infections | 7/13 (54%) |
| Immune dysregulation | 10/13 (77%) |
| ↓ % CD4 | 6/11 (55%) |
| Normal % CD4 subsets | 6/9 (67%) |
| Normal % Tregs | 5/9 (56%) |
| ↑ % CD8 | 6/11 (55%) |
| ↓ naïve, ↑ memory CD8 | 5/10 (50%) |
| ↓ % MAIT | 3/3 (100%) |
| Normal % CD19 | 5/8 (63%) |
| Autoantibodies | 6/10 (60%) |

### Table S5

| Target | Clone | Fluorophore | Catalogue # | Supplier |
|---|---|---|---|---|
| **Panel 1** | | | | |
| **Target** | **Clone** | **Fluorophore** | **Catalogue #** | **Supplier** |
| CD3 | UCHT1 | BV510 | 300448 | BioLegend |
| CD4 | SK3 | APC-R700 | 566909 | BD Biosciences |
| CD8 | RPA-T8 | BV421 | 562428 | BD Biosciences |
| CD25 | 2A3 | BV786 | 741035 | BD Biosciences |
| CD56 | NCAM16.2 | BUV661 | 750478 | BD Biosciences |
| CD127 | HIL-7R-M21 | PE-CF594 | 562397 | BD Biosciences |
| CD279 (PD-1) | EH12.1 | PE-Cy7 | 561272 | BD Biosciences |
| Helios | 22F6 | PE | 137216 | BioLegend |
| FOXP3 | 236A/E7 | AF488 | 561181 | BD Biosciences |
| IL-2 | MQ1-17H12 | BV650 | 564166 | BD Biosciences |
| IL-10 | JES3-19F1 | BB700 | 566568 | BD Biosciences |
| Fixable Viability | - | 575V | 565694 | BD Biosciences |
| **Panel 2** | | | | |
| **Target** | **Clone** | **Fluorophore** | **Catalogue #** | **Supplier** |
| CD3 | UCHT1 | BV510 | 300448 | BioLegend |
| CD4 | SK3 | BUV563 | 612912 | BD Biosciences |
| CD25 | 2A3 | BUV805 | 742070 | BD Biosciences |
| CD127 | HIL-7R-M21 | APC-R700 | 565185 | BD Biosciences |
| FOXP3 | 236A/E7 | AF488 | 561181 | BD Biosciences |
| Helios | 22F6 | PE | 137216 | BioLegend |
| IL-4 | MP4-25D2 | PE-CF594 | 565161 | BD Biosciences |
| IL-9 | MH9A3 | BV421 | 564254 | BD Biosciences |
| IL-17A | N49-653 | PerCP-Cy5.5 | 560799 | BD Biosciences |
| IL-21 | 3A3-N2.1 | AF647 | 560493 | BD Biosciences |
| IL-22 | 2G12A41 | PE-Cy7 | 366708 | BioLegend |
| IFN-$\gamma$ | 4S.B3 | BV711 | 564793 | BD Biosciences |
| TNF-$\alpha$ | MAb11 | BUV395 | 563996 | BD Biosciences |
| Fixable Viability | - | 575V | 565694 | BD Biosciences |
| **Panel 3** | | | | |
| **Target** | **Clone** | **Fluorophore** | **Catalogue #** | **Supplier** |
| CD3 | UCHT1 | BV510 | 300448 | BioLegend |
| CD4 | SK3 | APC-R700 | 566909 | BD Biosciences |
| CD8 | HIT8a | BV570 | 624298 | BD Biosciences |
| CD19 | HIB19 | BUV737 | 741829 | BD Biosciences |
| CD27 | O323 | BV605 | 751673 | BD Biosciences |
| CD38 | HIT2 | APC | 555462 | BD Biosciences |
| CD45RA | HI100 | BV421 | 562885 | BD Biosciences |
| IgD | IA6-2 | BUV615 | 613008 | BD Biosciences |
| IgM | G20-127 | BB515 | 564622 | BD Biosciences |
| IL-2 | MQ1-17H12 | BV650 | 564166 | BD Biosciences |
| IFN-$\gamma$ | 4S.B3 | BV711 | 564793 | BD Biosciences |
| TNF-$\alpha$ | MAb11 | BUV395 | 563996 | BD Biosciences |
| Helios | 22F6 | PE | 137216 | BioLegend |

### Table S5

| | | | | |
|---|---|---|---|---|
| Fixable Viability | - | 780 | 565388 | BD Biosciences |
| **Panel 4** | | | | |
| **Target** | **Clone** | **Fluorophore** | **Catalogue #** | **Supplier** |
| CD3 | SK7 | APC-H7 | 560176 | BD Biosciences |
| CD16 | 3G8 | BV510 | 563830 | BD Biosciences |
| CD27 | O323 | BV605 | 751673 | BD Biosciences |
| CD56 | NCAM16.2 | BUV661 | 750478 | BD Biosciences |
| CD57 | NK-1 | FITC | 555619 | BD Biosciences |
| CD94 | HP-3D9 | BUV805 | 748786 | BD Biosciences |
| Helios | 22F6 | PE | 137216 | BioLegend |
| IFN-γ | 4S.B3 | BV711 | 564793 | BD Biosciences |
| TNF-α | MAb11 | BUV395 | 563996 | BD Biosciences |
| Perforin | δG9 | PE-CF594 | 563763 | BD Biosciences |
| Granzyme B | GB11 | AF647 | 561999 | BD Biosciences |
| Fixable Viability | - | 575V | 565694 | BD Biosciences |
| **Panel 5** | | | | |
| **Target** | **Clone** | **Fluorophore** | **Catalogue #** | **Supplier** |
| CD4 | RPA-T4 | FITC | 555346 | BD Biosciences |
| CD25 | BC96 | APC | 567316 | BD Biosciences |
| CD127 | HIL-7R-M21 | BV421 | 562436 | BD Biosciences |
| Fixable Viability | - | 780 | 565388 | BD Biosciences |
| **Panel 6** | | | | |
| **Target** | **Clone** | **Fluorophore** | **Catalogue #** | **Supplier** |
| CD3 | UCHT1 | APC | 300412 | BioLegend |
| CD4 | OKT4 | PE | 317409 | BioLegend |
| CD8 | HIT8a | FITC | 300906 | BioLegend |
| CPD | - | eF450 | 65-0842-85 | ThermoFisher |
| Fixable Viability | - | eF780 | 65-0865-14 | ThermoFisher |
| **Panel 7** | | | | |
| **Target** | **Clone** | **Fluorophore** | **Catalogue #** | **Supplier** |
| CD4 | OKT4 | BV421 | 317434 | BioLegend |
| CD14 | 61D3 | APC-eF780 | 47-0149-42 | ThermoFisher |
| CD25 | 4E3 | PE | 130-113-282 | Miltenyi Biotec |
| CD127 | HIL-7R-M21 | R718 | 566967 | BD Biosciences |
| Fixable Viability | - | eF780 | 65-0865-14 | ThermoFisher |
