## Supplement_Text for "A germline heterozygous dominant negative *IKZF2* variant causing syndromic primary immune regulatory disorder and ICHAD"

950 West 28^th^ Avenue

Vancouver, BC, V5Z 4H4, Canada

**SUPPLEMENTAL METHODS**

**Sanger sequencing**

Genomic DNA was extracted from the whole blood of the patient and 2 healthy controls using a DNeasy Blood and Tissue Kit (Qiagen, Hilden, Germany). The breakpoint region was PCR amplified using GoTaq Green Master Mix (Promega, Madison, Wisconsin) using the following primers: Forward, 5’-GTACACTACACTACTCTGGA-3’ and Reverse, 5’-GTGGTTGAGTATGTTTCCTAG-3’ (Integrated DNA Technologies, Coralville, Iowa). The PCR product was purified using a QIAquick PCR Purification Kit (Qiagen) and sequenced using the same primers.

Plasmids were sequenced using the following primers:

| **Name** | **Sequence (5’-3’)** |
| --- | --- |
| Forward Primer 1 | ACGCTTACAATTTCCATTCGCCA |
| Forward Primer 2 | TGGCAGTACACCAATGGGCGTG |
| Forward Primer 3 | GGCCATGATGAGGGTAGCAGCC |
| Forward Primer 4 | GTCACCATGTACCTCCTATGGAAGA |
| Forward Primer 5 | GCAGCCATGATGACCACCAGTCC |
| Forward Primer 6 | GGAAGTTGCCACTCCAGTGCCC |
| Forward Primer 7 | ACCGGTGGGACATTTGAGTTGCT |
| Forward Primer 8 | TGTCATAGCTGTTTCCTGTGTGA |
| Forward Primer 9 | TTTCCCCCTGGAAGCTCCCTCG |
| Forward Primer 10 | TCCTTTGATCTTTTCTACGGGGT |
| Forward Primer 11 | TGCTACAGGCATCGTGGTGTCA |
| Forward Primer 12 | AGGGAATAAGGGCGACACGGAA |
| Forward Primer 13 | CCAAACTGGAACAACACTCAACCC |

**Helios plasmid cloning**

RNA was extracted from patient-derived and control LCLs using an RNeasy Plus Mini kit (#74034, Qiagen, Hilden, Germany) then converted to cDNA using the SuperScript III First-Strand Synthesis SuperMix (#11752050, ThermoFisher, Waltham, Massachusetts) according to manufacturer’s recommendations. Restriction sites flanking *IKZF2* cDNA were introduced using KpnI/XbaI primers with Phusion Plus DNA Polymerase (#F630S, ThermoFisher) according to manufacturer’s recommendations. The following primers were used: Forward, 5’-ATGCGGTACCGACGATGGAAACAGAGGCTATTGATG-3’ (KpnI) and Reverse, 5’-GACTTCTAGACTAGTGGAATGTGTGCTCCC-3’ (XbaI). PCR products were digested with KpnI and XbaI then cloned into a pCMV6-XL4-3xFLAG vector and confirmed by Sanger sequencing.

***IL2* promoter luciferase plasmid cloning**

XhoI and HindIII restriction sites were introduced to flank the human *IL2* gene promoter fragment spanning -580 base pairs (bp) upstream to +20 bp downstream of the transcription start site from HEK293 genomic DNA by PCR. The following primers were used: Forward, 5’-CCGTCTCGAGACAGTACCTCAAGCTCAATAAGCA-3’ (XhoI) and Reverse, 5’-CCCTAAGCTTAAAGAGAGTGATAGGGAACTCTTGA-3’ (HindIII). PCR products were digested with XhoI and HindIII and subsequently cloned into the pGL4.14 firefly luciferase gene reporter vector (Promega, Madison, Wisconsin) to generate a pIL2-Luc2 plasmid.

**Transient plasmid transfections**

For immunoblotting and immunofluorescence, HEK293 cells were seeded at 8.0x10^5^ cells/well in a 6-well plate in complete DMEM supplemented with 10% FBS (Gibco; Life Technologies, Carlsbad, California) and incubated for 24h at 37^0^C. Cells were transfected with 3μg of plasmid DNA using the P3000, Lipofectamine 3000, and Opti-MEM Medium reagents according to manufacturer’s recommendations. For luciferase transfections, HEK293 cells were seeded at 1.5x10^5^ cells/well in 24-well plates in complete DMEM supplemented with 10% FBS and incubated for 24 h at 37^0^C. Cells were then transfected with 250ng of pIL2-Luc2, 250ng of EV, WT, or p.Gly136_Val192dup Helios, and 10ng Renilla luciferase (R-Luc) using Lipofectamine 3000 as above. Transfected cells were then lysed and processed using a Dual-Glo Luciferase Assay Kit (#E2920, Promega). Luciferase activity was read on a Tecan Infinite M200 plate reader (Tecan, Männedorf, Switzerland). For immunoblotting, immunofluorescence, and luciferase assays, variant/WT ratios were varied by maintaining the total plasmid DNA amount the same (e.g. 1:1 p.Gly136_Val192dup/WT was 1.5μg variant + 1.5μg WT).

**Luciferase assay analyses**

Firefly luciferase relative light units (RLU) were normalized to renilla luciferase RLU (firefly/renilla). *IL2* promoter activity was defined as normalized firefly RLU relative to EV (condition/EV).

**Cell culture and lymphoblastoid cell line (LCL) immortalization**

PBMCs from the patient and controls were cultured or stimulated in complete RPMI-1640 (GE Healthcare) supplemented with 10% heat inactivated FBS (Gibco, Life Technologies, Rockville, Maryland), 2mM _L_-glutamine (HyClone; ThermoFisher Scientific, Waltham, Massachusetts), and 1mM sodium pyruvate (Gibco; Life Technologies, Carlsbad, California).

To generate Epstein-Barr virus (EBV)-transformed immortalized B cell lines, PBMCs from the patient and a healthy control were cultured in complete RPMI-1640 supplemented with tacrolimus (AG Scientific, San Diego, California) for 1h. Cells were then infected with supernatant collected from the viral replication-permissive marmoset cell line B95-8 (ATCC, Manassas, Virginia) until sufficient B cell blasts were observed. LCLs were cultured in complete RPMI-1640 without tacrolimus.

**Immunoblotting**

Whole cell lysates were prepared by lysing LCLs, THP-1, Jurkat T cells, and transfected HEK293s in a RIPA Lysis and Extraction Buffer (#89901, ThermoFisher) supplemented with Halt Protease and Phosphatase Inhibitor Cocktail (#78440, ThermoFisher). Lysates were separated by 10% SDS-PAGE, transferred onto polyvinylidene difluoride membranes (Immobilon-FL; MilliporeSigma, Billerica, Massachusetts), blocked with 5% bovine serum albumin (BSA) in Tris-buffered saline with Tween-20, incubated with primary antibodies for 18h at 4^0^C, incubated with secondary antibodies for 1h at room temperature, and imaged on a LI-COR Odyssey infrared scanner (LI-COR Biosciences, Lincoln, Nebraska). The following primary antibodies were used: Helios (#89270S) and β-actin (#8457S, #3700S), all from Cell Signaling Technologies (Danvers, Massachusetts), and FLAG (clone M2, #F3165, Millipore Sigma). The secondary antibodies used were the following: goat anti-rabbit IgG Dylight 800 conjugated (#611-145-002-0.5, Rockland Immunochemicals, Pottstown, Pennsylvania) and goat anti-mouse IgG IRDye 680RD (#926-6870, LI-COR). Immunoblots were quantified using Image Studio Lite (v.5.2.5, LI-COR).

**Immunofluorescence**

12mm coverslips (Fisherbrand, Fisher Scientific, Waltham, Massachusetts) were sterilized with 70% ethanol and washed with PBS. Sterilized coverslips were placed in 6-well plates before HEK293 cells were seeded and transfected as described above. Cells were subsequently fixed with 4% paraformaldehyde in PBS (#15710, Electron Microscopy Sciences, Hatfield, Pennsylvania) for 10min, washed with PBS, permeabilized with 0.01% Triton X-100 in PBS, and washed with PBS. Transfected cells were stained with Helios primary antibodies (#720419, Invitrogen, ThermoFisher), washed, stained with goat anti-rabbit IgG conjugated to Alexa Fluor 647 (#A32733, ThermoFisher) and Alexa Fluor 488 Phalloidin (#A12379, ThermoFisher), then washed. Coverslips were mounted in ProLong Gold Antifade Mountant with DAPI (#P36935, ThermoFisher) on Superfrost Plus Microscope Slides (Fisherbrand, Fisher Scientific). Slides were imaged on a Leica SP5 II laser Scanning Confocal Microscope (Leica Microsystems GmbH, Wetzlar, Germany). Images were analyzed using Fiji (NIH, Bethesda, Maryland).

**Immunophenotyping and analysis**

PBMCs from the patient or controls were thawed and stimulated with 50ng/mL of phorbol 12-myristate 13-acetate (PMA) and 1μM ionomycin (P/I) for 4h in the presence of GolgiStop (#554724, BD Biosciences, Franklin Lakes, New Jersey). Cells were stained with extracellular antibodies from panels 1-4 (**Supplemental Table 5**) then fixed and permeabilized using the eBioscience Foxp3 Transcription Factor Staining Buffer Set (#00-5523-00, Invitrogen, ThermoFisher) according to manufacturer’s recommendations. Samples were subsequently stained with intracellular antibodies from panels 1-4 (**Supplemental Table 5**) and acquired on a FACSymphony flow cytometer (BD Biosciences). Compensation and voltages were determined according to single stain controls using species-specific Compensation Particles for antibodies (#552843, #552845, #552844, BD Biosciences) and ArC Amine Reactive Compensation Beads for viability dyes (#A10346, ThermoFisher). Gating, frequency, and expression analyses were carried out on FlowJo (BD Biosciences).

NK cell clustering analyses were carried out by gating on CD3^-^CD56^+^ NK cells from the patient and controls, exporting the NK cell populations, then concatenating the unstimulated and stimulated conditions from all individuals. The concatenated unstimulated and stimulated samples were clustered on CD16, CD27, CD57, CD94, IFN-γ, TNF-α, granzyme B, perforin, and Helios using the tSNE plugin, opt-SNE^1^, 1000 iterations, perplexity at 30, learning rate at 18925, KNN algorithm set to exact (vantage point tree), and using the Barnes-Hut gradient algorithm.

**Proliferation assays and analysis**

Thawed PBMCs were seeded at 25,000, 50,000, or 100,000 cells/well in technical triplicate in 96-well plates and labelled with Cell Proliferation Dye eF450 (#65-0842-85, Invitrogen, ThermoFisher) for 10min at 37^0^C in the dark. Cell labelling was stopped with cold complete RPMI-1640 for 5min at 4^0^C, washed, then stimulated with 1:8, 1:16, 1:32 bead to PBMC ratios using anti-CD3/CD28-coated Dynabeads (#11141D, Human T-Expander, Gibco, ThermoFisher) for four days. Cells were stained with antibody panel 6 (**Supplemental Table 5**), acquired on a CytoFLEX (Beckman Coulter), and analyzed using FlowJo. Percent divided cells were quantified using the Proliferation Modelling plugin on FlowJo and peak fitting.

**Treg and Tconv expansion**

CD4^+^ T cells were enriched from thawed patient or control PBMCs using a EasySep Human CD4^+^ T Cell Enrichment Kit (#19052, Stemcell Technologies, Vancouver, BC). CD4^+^ T cells were stained with panel 7 and sorted for CD4^+^CD25^+^CD127^lo^ Tregs and CD4^+^CD25^-^CD127^+^ Tconvs on an Astrios (Beckman Coulter, Brea, California) or FACS Aria (BD Bioscience) cell sorter. Tregs and Tconvs were plated on aAPCs, which are irradiated (75Gy) mouse L cells overexpressing hCD32, hCD58, hCD80 and loaded with 0.1μg/mL anti-CD3 (OKT3, University of British Columbia Antibody Lab, Vancouver, British Columbia). Tregs and Tconvs were cultured in complete X-VIVO 15 (#BE02-060Q, Lonza, Basel, Switzerland) supplemented with 5% human serum (#022-210, WISENT, St-Bruno, Quebec), 1% penicillin/streptomycin (Gibco, ThermoFisher), 2mM GlutaMAX (Gibco, ThermoFisher), and 15.97mg/L phenol red (Sigma-Aldrich, Burlington, Massachusetts). Tregs and Tconvs were expanded in X-VIVO 15 additionally supplemented with 1000 IU/mL IL-2 for Tregs and 100 IU/mL IL-2 for Tconvs (Proleukin, #02130181, Novartis, Basel, Switzerland) for seven days. IL-2 was refreshed every 2-3 days. Tregs and Tconvs were harvested on day 7 and rested in X-VIVO 15 supplemented with reduced IL-2 (100 IU/mL) for Tregs and no IL-2 for Tconvs. Tregs and Tconvs were then allocated for T cell suppression assays, cytokine quantitation, and TSDR methylation.

**T cell suppression assays**

CD3^+^ Tresp cells were enriched from thawed patient or control PBMCs using an EasySep Human T cell Isolation Kit (#17951, Stemcell Technologies). CD3^+^ Tresp cells were labelled and stimulated as described in the ‘Proliferation Assay’ section and cocultured with expanded patient or control Tregs and Tconvs for four days. As positive control, CD3^+^ Tresp cells were stimulated with beads alone. All CD3^+^ Tresp cells were plated at a density of 30,000 cells/well. CD4^+^ and CD8^+^ T cell proliferation was calculated by determining the division index (DI) using proliferation modelling as described above. Percent suppression was calculated using the following formula: (1 – [DI of sample/DI of positive control]) x 100%.

**Measuring Treg and Tconv cytokine production by LEGENDplex**

Supernatant was collected from the following conditions: 1) expanded Treg and Tconv cells were seeded at 25,000 cells/well and stimulated with 100U/mL of IL-2 and a 1:1 anti-CD3/CD28-coated Dynabeads to cell ratio for four days; 2) co-cultured Tregs and Tresp from suppression assays; and 3) PBMCs stimulated for proliferation. Supernatants were harvested and analyzed using a LEGENDplex Human Th Cytokine Panel 12-plex (#741027, BioLegend) on a CytoFLEX (Beckman Coulter) and analyzed with Qognit software (BioLegend) according to manufacturer’s recommendations.

**TSDR methylation**

Genomic DNA was isolated from expanded Tregs and Tconvs and bisulfite converted with an EZ DNA Methylation-Direct Kit (Zymo Research, Irvine, California). Samples were PCR amplified with the following primers: Forward, 5’-AGAAATTTGTGGGGTGGGGTAT-3’ and Reverse, 5’-ATCTACATCTAAACCCTATTATCACAACC-3’. Pyrosequencing was performed using a PyroMark Q96 MD (Qiagen) using the following primer: 5’-AGAAATTTGTGGGGTGGG-3’.

**Single-cell RNA sequencing of Tregs and Tconvs**

Patient and age-matched and sex-matched control PBMCs were thawed and enriched for CD3^+^ T cells using an EasySep Human T cell Isolation Kit (#17951, Stemcell Technologies). CD3^+^ T cells were stained with panel 5 (**Supplemental Table 5**) and sorted for CD4^+^CD25^+^CD127^lo^ Tregs and CD4^+^CD25^-^CD127^+^ Tconvs on a FACS Aria cell sorter (BD Biosciences) (**Supplemental Figure 7A**). Sorted cells were left unstimulated or stimulated with P/I for 4h. scRNA-seq sample preparation was performed following the BD Rhapsody Single Cell platform as previously described^2^ using the Rhapsody Cartridge Reagent Kit (#633731), the Rhapsody Cartridge Kit (#633733), the Rhapsody cDNA Kit (#633733), the Human Single Cell Sample Multiplexing Kit (#633781), and the Whole Transcriptome Analysis (WTA) Amplification Kit (#633801) all from BD Biosciences. Briefly, cells from each sample were labelled with Sample Tags and CD279 (PD-1) AbSeq (#940015, clone EH12.1, barcode sequence: ATGGTAGTATCACGACGTAGTAGGGTAATTGGCAGT), CD45RA AbSeq (#940011, clone HI100, barcode sequence: AAGCGATTGCGAAGGGTTAGTCAGTACGTTATGTTG), and HLA-DR AbSeq (#940010, clone G46-6, barcode sequence: TGTTGGTTATTCGTTAGTGCATCCGTTTGGGCGTGG) Ab-Oligos (all from BD Biosciences) and pooled in cold sample buffer to obtain ~20,000 cells in 620mL for each of the unstimulated and stimulated samples. A nanowell cartridge was primed and loaded with pooled samples for 15min at room temperature using the Rhapsody Express instrument (#633702, BD Biosciences) then loaded with cell capture beads. The cartridge was washed, cells were lysed, beads were retrieved, and the sample underwent reverse transcription, exonuclease I treatment, and denaturation. WTA, AbSeq, and Sample Tag libraries were prepared using a combination of random priming and extension and a series of PCR steps. PCR products were purified using AMPure beads (#A63880, Beckman Coulter) and quality was checked using an Agilent DNA High Sensitivity Kit (#5067-4626, Agilent Technologies, Santa Clara, California) on a 2100 Bioanalyzer (Agilent Technologies). Libraries were pooled according to manufacturer’s recommendations, diluted to 650pM with a 20% PhiX spike-in, and multiplexed for paired-end (2 x 115bp) sequencing on a NextSeq 2000 Sequencing System (Illumina) at 200 cycles P3 for 1.1 billion reads.

**scRNA-seq bioinformatics analyses**

FASTQ files were processed using the BD Rhapsody WTA Analysis Pipeline on SevenBridges ([www.sevenbridges.com](http://www.sevenbridges.com)) according to manufacturer’s recommendations, using the GRCh38 GENCODE v.29 reference genome and transcriptome annotation. Distribution-based error correction (DBEC)-adjusted molecule counts and the R package Seurat^3^ was used for all downstream analyses as previously described^2^. Tregs and Tconvs were scaled and clustered separately. Cell identities were annotated with the SingleR package using fine labeling from the Database of Immune Cell Expression, Expression quantitative trait loci (eQTLs) and Epigenomics project (DICE)^4^. CD45RA AbSeq and/or HLA-DR AbSeq distributions were used to define different cell subsets, including CD45RA^-^HLA-DR^+^ Tregs, CD45RA^+^HLA-DR^-^ Tregs, CD45RA^-^HLA-DR^-^ Tregs, and CD45RA^+^ naïve Tconvs and CD45RA^-^ memory Tconvs. Differential gene expression analyses were accomplished using the Seurat negative binomial regression^5^ as previously described^2^. Gene set enrichment was carried out using Enrichr^6^ with genes that were significantly (FDR<0.05) different between comparisons.

**SUPPLEMENTAL TABLE LEGENDS**

**Supplemental Table 1. Differentially expressed genes in Treg subsets.** Genes that had an adjusted p-value<0.05 when comparing P1 to controls in unstimulated or stimulated CD45RA^+^HLA-DR^-^, CD45RA^-^HLA-DR^+^, CD45RA^-^HLA-DR^-^ Tregs. Shown are gene names, average log_2_(fold change), percent reads in the patient or controls, and adjusted p-values.

**Supplemental Table 2. Differentially expressed genes in Tconv subsets.** Genes that had an adjusted p-value<0.05 when comparing P1 to controls in unstimulated or stimulated CD45RA^+^ naïve or CD45RA^-^ memory CD4^+^ Tconvs. Shown are gene names, average log_2_(fold change), percent reads in the patient or controls, and adjusted p-values.

**Supplemental Table 3. Major clinical and laboratory features associated with pathogenic germline Helios variants.** Tabulation of clinical and immunological features observed in all patients with pathogenic *IKZF2* variants described to date. Variants are ordered from the N-terminus to the C-terminus of Helios. Asterisks represent variants that have not been tested in the heterozygous state to assess whether they are dominant negative. Horizontal lines represent features that were not present. Het., heterozygous; LOF, loss-of-function; DN, dominant negative; ND, no data; HLH, hemophagocytic lymphohistiocytosis; AIHA, autoimmune hemolytic anemia; ITP, idiopathic thrombocytopenic purpura; SLE, systemic lupus erythematosus; EBV, Epstein-Barr virus; EM, effector memory; CM, central memory; Treg, regulatory T cells; NK, natural killer cells; IVIG, intravenous immunoglobulin replacement.

**Supplemental Table 4. Red flags of germline Helios deficiency.** Tabulation of the most frequently (>50% of patients) observed genetic, clinical, and laboratory features in patients.

**Supplemental Table 5. Flow cytometry antibodies used.** List of flow cytometry antibodies used to identify different cell subsets organized based on panel.

**SUPPLEMENTAL FIGURE LEGENDS**

**Supplemental Figure 1. Extended NK cell phenotyping.** A-B) Representative histograms of A) CD56 and B) CD16 in P1 and control CD3^-^CD56^+^ NK cells. Indicated is how CD56^dim^, CD56^bright^, CD16^-^, and CD16^+^ NK cells are defined as well as their associated frequencies. C-G) Representative contour plots of C) CD57^+^, D) CD27^+^, E) CD94^+^, F) CD8^+^, and G) CD25^+^ NK cells in P1 and a control. H-N) Quantification of H) CD16^+^, I) CD57^+^, J) CD27^+^, K) CD94^+^, L) CD8^+^, M) CD25^+^, N) Helios mean fluorescence intensity (MFI) in CD56^bright^ and CD56^dim^ NK cells. O-P) Representative contour plots of O) IFN-γ^+^ and P) TNF-α^+^ NK cells in P1 and a control. Q-R) Quantification of Q) granzyme B and R) perforin MFI from CD56^bright^ and CD56^dim^ NK cells. S) Clustering of live NK cells stimulated with PMA+ionomycin. Each panel is coloured based on marker expression. H-K), N), Q-R) P1 n=5, HC^A^ (adult control) n=8, HC^P^ (pediatric control) n=11. L-M) P1 n=3, HC n=8. *p<0.05, **p<0.01, ***p<0.001, ****p<0.0001. Ordinary one-way ANOVA with Šidák’s multiple comparisons test.

**Supplemental Figure 2. Patient B cell immunophenotyping.** A) Representative dot plots for CD3^-^CD19^+^ B cells in P1 and a control. B) Quantification of A). C) Representative contour plots for IgD^+^CD27^-^ naïve (NB), IgD^+^CD27^+^ non-switched memory (NSM), and IgD^-^CD27^+^ switched memory (SM) B cells from P1 and a control. Quadrants corresponding to each subset are shown to the right. Frequencies are indicated. D-F) Quantification of C) for D) NB, E) NSM, and F) SM B cell frequencies. G) Representative contour plots for IgM^hi^CD38^hi^ transitional B cells (TrB) from P1 and a control. H) Quantification of G). I) Representative contour plots for CD27^+^CD38^+^ plasmablasts (PB). J) Quantification of I). K) Representative Helios histograms for CD19^+^ B cells in P1 and controls compared to a fluorescence minus one (FMO) control. Mean fluorescence intensities (MFI) are indicated. L) Quantification of Helios MFI in different B cell subsets. M) Representative contour plots of TNF-α^+^ B cells in P1 and a control. N-P) Quantification of TNF-α^+^ N) NSM, O) SM, and P) NB cells in P1 and controls. A-F), H), J-L), N-P) P1 n=5, HC^A^ (adult control) n=8, HC^P^ (pediatric control) n=11. *p<0.05, **p<0.01, ***p<0.001, ****p<0.0001. Ordinary one-way ANOVA with Šidák’s multiple comparisons test.

**Supplemental Figure 3. Extended T_H_ phenotyping.** A) Representative contour plots for T_H_17 from P1 and a control. Quadrants corresponding to each subset are indicated. Frequencies are included. B) Quantification of T_H_17 cells. C-N) PBMCs stimulated with anti-CD3/CD28 beads at 1:8, 1:16, and 1:32 bead to cell ratios for 4 days. Cytokine concentrations measured by LEGENDplex for C) IL-2, D) IL-4, E) IL-5, F) IL-6, G) IL-9, H) IL-10, I) IL-13, J) IL-17F, K) IL-17A, L) IL-22, M) TNF-α, and N) IFN-γ. Error bars represent technical duplicates. A-B) P1 n=5, HC^A^ (adult control) n=8, HC^P^ (pediatric control) n=11. *p<0.05, **p<0.01. Ordinary one-way ANOVA with Šidák’s multiple comparisons test.

**Supplemental Figure 4. Extended Treg phenotyping.** A) Gating strategy for identifying CD3^+^CD4^+^CD25^+^CD127^lo^FOXP3^+^ Tregs. B) Average Treg-specific demethylation region (TSDR) methylation across CpG sites. n=2. C-D) Representative contour plots for C) PD-1^+^ and D) IL-9^+^ and IL-21^+^ Tregs from P1 and a control. E-I) Isolated and expanded Tregs or Tconv stimulated with anti-CD3/CD28 beads at 1:1 bead to cell ratio for 4 days. Cytokine concentrations were measured by LEGENDplex for E) IL-4, F) IL-5, G) IL-6, H) IL-9, I) IL-10, J) IL-13, K) IL-17A, L) IL-17F, M) IL-22, N) TNF-α, and O) IFN-γ. Shown are technical triplicates.

**Supplemental Figure 5. P1 Tresp cytokine production in presence of control Tregs.** A-L) P1 CD3^+^ Tresp cells stimulated with anti-CD3/CD28 beads and cocultured with control Tregs. Cytokine concentrations were measured by LEGENDplex for A) IL-2, B) IL-4, C) IL-5, D) IL-6, E) IL-9, F) IL-10, G) IL-13, H) IL-17F, I) IL-17A, J) IL-22, K) TNF-α, and L) IFN-γ. Shown are technical triplicates.

**Supplemental Figure 6. scRNA-seq workflow and extended analyses.** A) General workflow for the preparation of samples for scRNA-seq. B-F) Volcano plots comparing unstimulated P1 and control B) CD45RA^+^HLA-DR^-^ naïve Tregs, C) CD45RA^-^HLA-DR^+^ activated Tregs, D) CD45RA^-^HLA-DR^-^ Tregs, E) CD45RA^+^ naïve Tconvs, and F) CD45RA^-^ memory Tconvs. Red=significantly increased, blue=significantly decreased, gray=non-significant. Vertical dashed line: fold change=1.5, horizontal dashed line: FDR=0.05. Top 10 significant genes are labelled. G) Gene set enrichment of differentially expressed genes between unstimulated naïve patient and control Tconvs using EnrichR. Shown are the combined scores and adjusted p-values. H=MSigDB Hallmark, B=BioPlanet 2019, N=NCI Nature 2016, K=KEGG 2021 Human.

**Supplemental Figure 7. Location and impact of human germline *IKZF2* variants.** A) Schematic representation of the Helios protein and domains. Annotated are all germline *IKZF2* variants identified to date. The variant included in the current study is marked in red. B) Moving slider model of impact of germline *IKZF2* variants on Helios function ranging from loss-of-function (LOF) to hypomorphic to wild-type (WT) to hypermorphic to gain-of-function (GOF). Variants have been subjectively placed on the slider based on their estimated/reported impact on Helios function.
